# Multi-ancestry meta-analysis of host genetic susceptibility to tuberculosis identifies shared genetic architecture

**DOI:** 10.1101/2022.08.26.22279009

**Authors:** Haiko Schurz, Vivek Naranbhai, Tom A. Yates, James J. Gilchrist, Tom Parks, Peter J. Dodd, Marlo Möller, Eileen G Hoal, Andrew P. Morris, Adrian V.S. Hill, the International Tuberculosis Host Genetics Consortium

## Abstract

The heritability of susceptibility to tuberculosis disease (TB) has been well recognized. Over one-hundred genes have been studied as candidates for TB susceptibility, and several variants were identified by genome-wide association studies (GWAS), but few replicate. We established the International Tuberculosis Host Genetics Consortium (ITHGC) to perform a multi-ancestry meta-analysis of GWAS including 14153 cases and 19536 controls of African, Asian, and European ancestry. Our analyses demonstrate a substantial degree of heritability (pooled polygenic h^2^=26.3% 95% CI 23.7-29.0%) for susceptibility to TB that is shared across ancestries, highlighting an important host genetic influence on disease. We identified one global host genetic correlate for TB at genome-wide significance (p<5×10^−8^) in the human leukocyte antigen (HLA)-II region (rs28383206, p-value = 5.2×10^−9^). These data demonstrate the complex shared genetic architecture of susceptibility to TB and the importance of large scale GWAS analysis across multiple ancestries experiencing different levels of infection pressures.

## Introduction

Tuberculosis (TB), caused by *Mycobacterium tuberculosis (Mtb)* and related species, remains a leading cause of death globally. Around one-quarter of the global population is estimated to show immunological evidence of prior exposure to *Mtb*^1^, and in 2019 an estimated 10 million people developed disease resulting in 1.4 million deaths^2^. This disease burden could be substantially reduced with action to address the social determinants of disease and equitable scale-up of existing interventions. However, tools to prevent, diagnose and treat TB could be improved if a better understanding of the underpinning pathophysiology could help identify those at greatest risk of disease.

The role of host genetic factors in TB susceptibility has long been of significant interest. Over one-hundred candidate genes have been studied, but few associations have proven reproducible^3^. This failure to replicate may be a result of the modest size of many TB genome-wide association studies (GWAS), variability in phenotyping between studies, the impact of population specific effects, the challenge of complex population structure in some high burden settings (e.g. admixed individuals) and, possibly, pathogen variation^4–10^. Seventeen GWAS have been reported but only two loci replicate between studies^5,10–25^. The *WT1* locus, identified in cohorts from Ghana and Gambia, replicated in South Africa and Russia. The *ASAP1* locus identified in Russia was replicated through re-analysis of prior studies^4,7^.

To address these challenges, we established the International Tuberculosis Host Genetics Consortium (ITHGC) to study the host genetics of disease through collaborative and equitable data sharing^3^. The ITHGC includes 12 case-control GWAS studies from 9 countries in Europe, Africa, and Asia (total of 14153 pulmonary TB cases and 19536 healthy controls). Inclusion of multiple ancestral groups in a multi-ancestry meta-analysis has the advantage of maximizing power and enhancing fine-mapping resolution to identifying true global associated variants that influence TB susceptibility across population groups.

Here we present the first analyses of the ITHGC dataset exploring host genetic correlates of TB susceptibility using a multi-ancestry meta-analysis approach, including fine mapping of HLA loci and estimation of genetic heritability.

## Results

### Study overview

In total 12 GWAS from three major ancestral groups (European, African, and Asian) were included in this study (Table 1), a more detailed table outlining selection of cases and controls is provided in Table S1. All individual datasets were imputed and aligned to the same reference allele before association testing, using an additive genetic model, to obtain odds ratios (OR) and p-values to be used in the meta-analysis. For each individual study (for which we had raw genotyping data) the polygenic heritability was estimated, and HLA alleles were imputed for fine-mapping of the HLA regions.

**Table 1:**
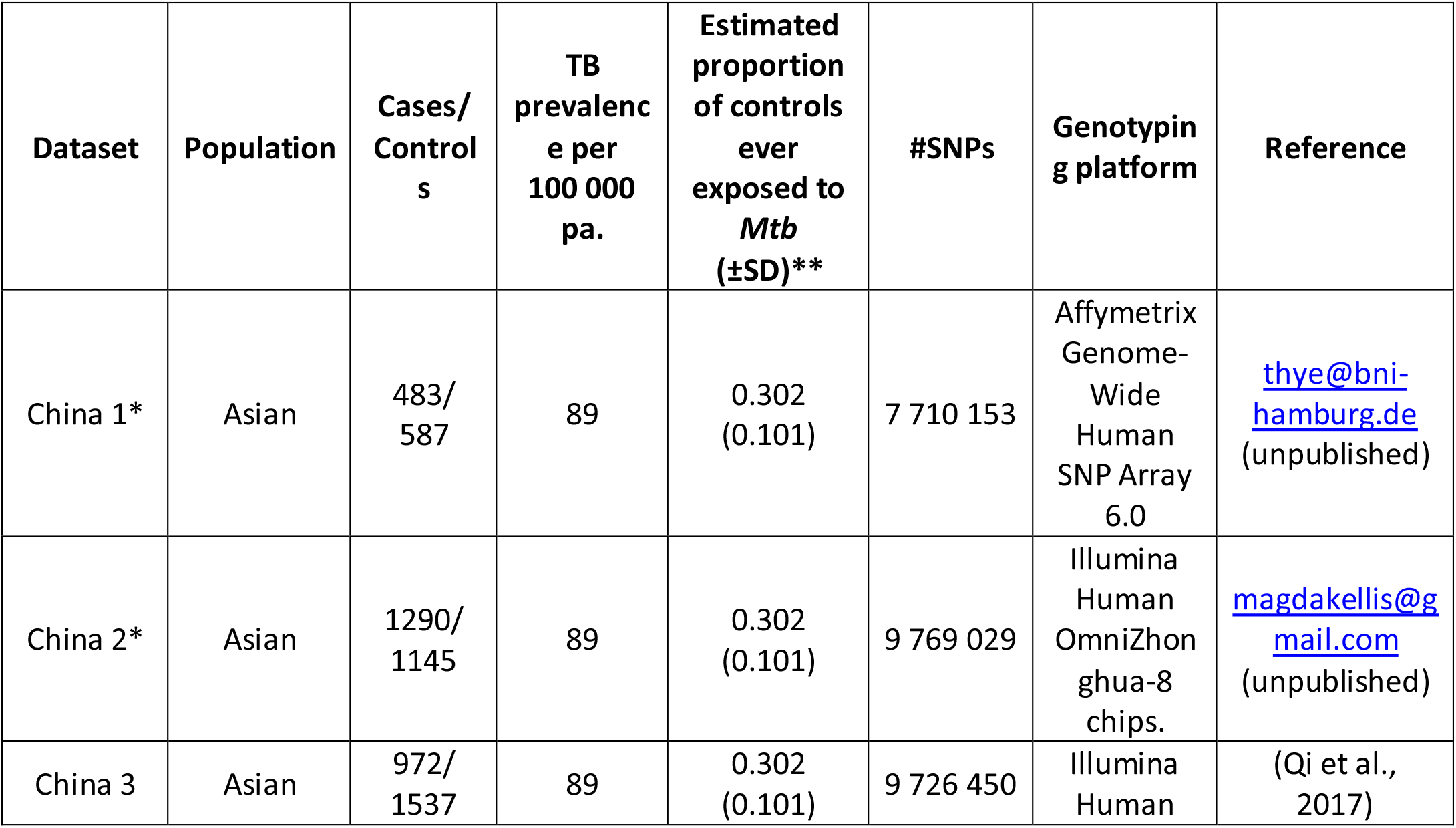
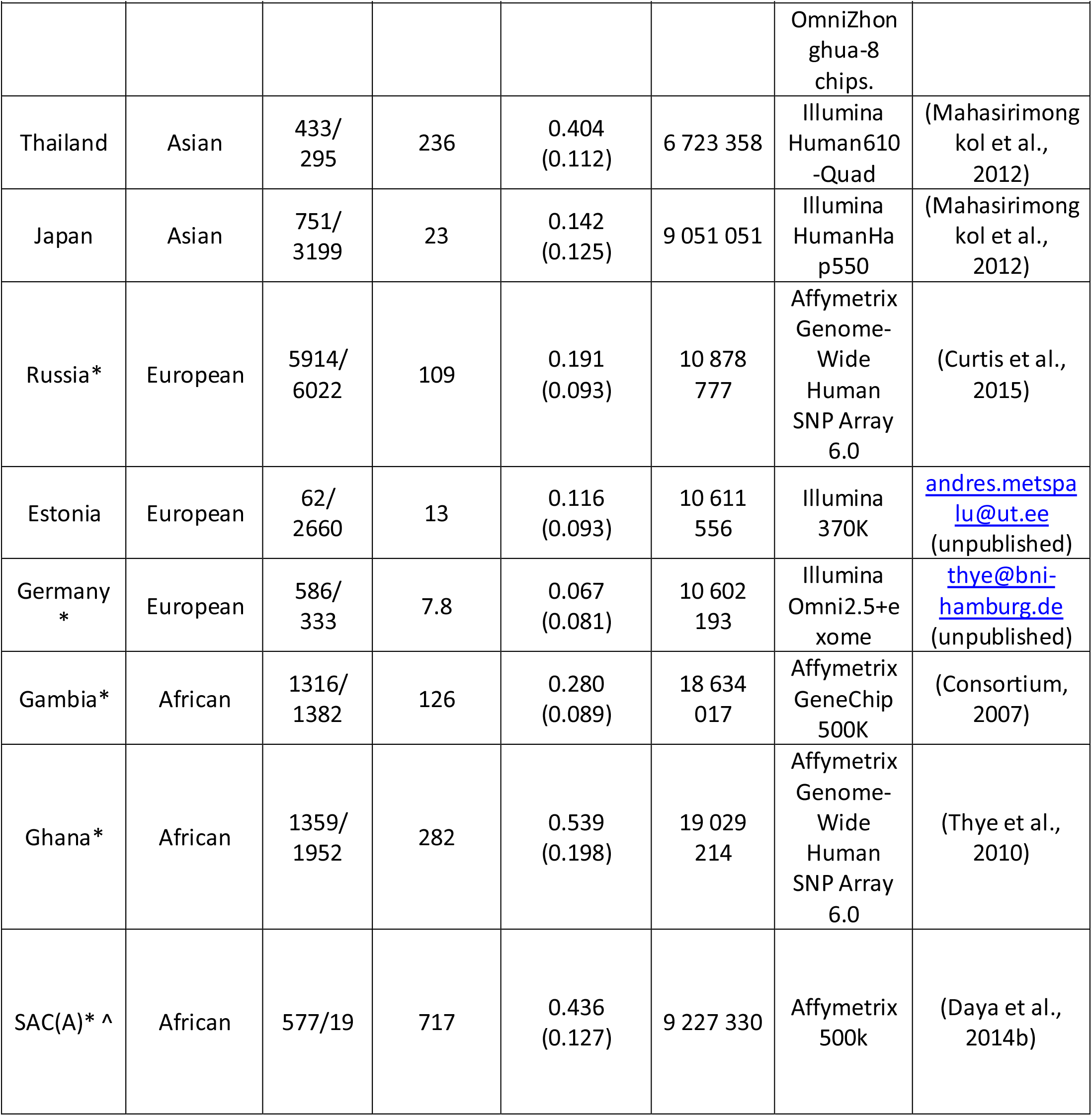
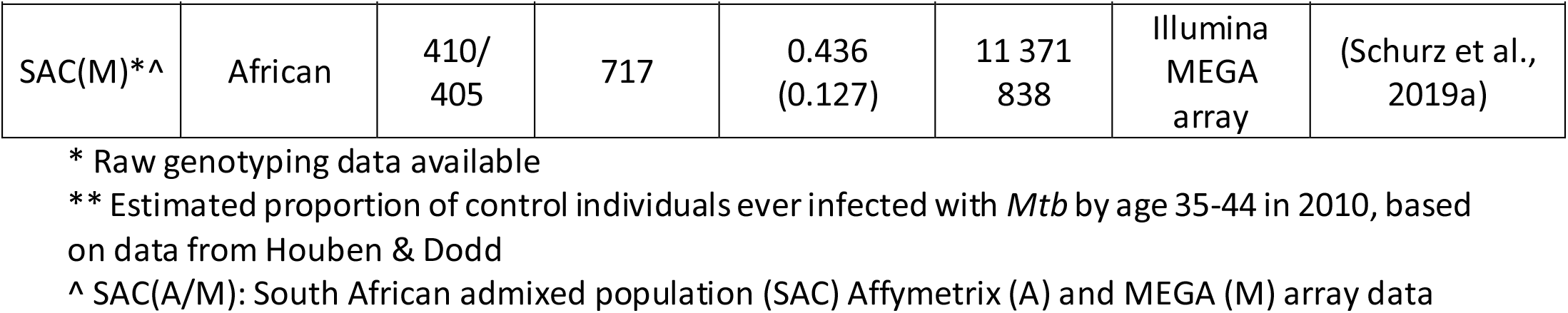
Summary of ITHGC TB-GWAS datasets.

The summary statistics from the individual GWAS of each dataset were used to conduct a combined, multi-ancestry meta-analysis using MR-MEGA and ancestry-specific (European, African, and Asian) fixed effects (FE) meta-analyses using GWAMA. Finally, the impact of infection pressure on the multi-ancestry meta-regression was assessed and the concordance in direction of effect for the reference allele between studies was investigated.

### Polygenic heritability estimates suggest a genetic contribution to susceptibility

Twin studies estimate the narrow sense heritability of susceptibility to TB at up to 80% (Diehl and von Verschner 1933, Kallman and Reisner 1943, Comstock 1978), but there are few modern estimates. Using raw (unimputed) genotyping data, and assuming population prevalence of disease in each study population equivalent to the reported WHO prevalence rates for that country^2^, we estimated polygenic heritability of susceptibility to TB in ten contributing studies which ranged from 5-36% (average of 26.3%, Table S2). Comparisons of the heritability estimates between studies from different geographical locations do not take into consideration differences in environmental pressures between the included studies and as such these estimates of heritability are only interpretable if the distribution of non-genetic determinants of TB are held constant^26^. Recent history has seen the near elimination of TB in several countries associated with economic development and public health action. However, while improvement of socioeconomic standing and environment have a stronger impact than host genetics, these crude estimates of polygenic heritability do indicate that TB susceptibility is, in part, heritable. These results require future, more rigorous investigations to narrow down the level of heritable risk and pinpoint genomic loci involved by accounting for population stratification to obtain more accurate heritability estimates.

### Multi-ancestry meta-analysis identifies new susceptibility loci for TB

For the primary multi-ancestry meta-analysis MR-MEGA was used as it allows for differences in allelic effects of variants on disease risk between GWAS. Principal components (PC), derived from a matrix of similarities in allele frequencies between GWAS, were plotted and revealed distinct separation between the three main ancestral groups included in the study (Figure 5). To account for this the first two PCs were included as covariates in MR-MEGA as they sufficiently accounted for the allele frequency differences between the study populations, as assessed via a QQ-plot and associated lambda inflation value (Figure S1, lambda = 1.00). In total 26,620,804 variants with a MAF >1% and present in at least three studies were included in the analysis, of which 3,184,478 were present in all 12 datasets.

After removing variants that displayed within ancestry heterogeneity (Cochran’s-Q p-value <0.1 in Europe, Asia or Africa) and associations that were driven by a single study, a significant peak on chromosome 6 was identified in the *HLA class ll* region (Figure 2). One variant (rs28383206) within this peak was associated with susceptibility to TB at genome-wide significance (p<5×10^−8^, Figure 3 and 4, Table 2). Both the residual heterogeneity (p-value = 0.012) and ancestry-correlated heterogeneity (p-value = 5.28e^-6^) are significant (p-value < 0.05) for the associated variant. However, the evidence of ancestry-correlated heterogeneity is much stronger than for residual heterogeneity, indicating that genetic ancestry contributes more to differences in effects sizes between GWAS than does study design (phenotyping differences etc.). The association peak encompasses many *HLA-ll* genes, including *HLA-DRB1/5* (major histocompatibility complex, class II, DR beta 1/5), *HLA-DQA1* (major histocompatibility complex, class II, DQ alpha 1) and *HLA-DQB3* (major histocompatibility complex, class II, DQ beta 3, Figure 3). While not reaching genome-wide significance, the HLA class l locus is also indirectly tagged through the association with rs2621322, in the *TAP2* (transporter 2, ATP binding cassette subfamily B member) gene, a transporter protein that restores surface expression of MHC class I molecules and has previously been implicated in TB susceptibility^27^. *HLA-A, DQA1, DQB1, DRB1* and *TAP2* genes have previously been linked to TB susceptibility through TB candidate gene and GWAS analysis^27–31^. The HLA-II locus encodes several proteins crucial in antigen presentation including HLA-DR, HLA-DQ and HLA-DP, which are widely implicated in susceptibility to infection and autoimmunity^32,33^.

**Table 2:** Significant and suggestive associations (p-value <= 1e^-5^) for the multi-ancestry analysis including data from all 12 datasets implementing MR-Mega analysis with GCC.

| Marker Name | Chromosome | Position | Gene | Location | CADD score | EA^ | NEA^ | EAF^ | Sample size | Datasets | P-value |
| --- | --- | --- | --- | --- | --- | --- | --- | --- | --- | --- | --- |
| rs28383206 | 6 | 32575167 | *HLA-DRB1* | intergenic | 7.6 | G | A | 0.168 | 25059 | 8 | $8.26e^{-09}$ |

^EA and EAF: Effect allele and effect allele frequency
^NEA: Non-effect allele

**Figure 1:**
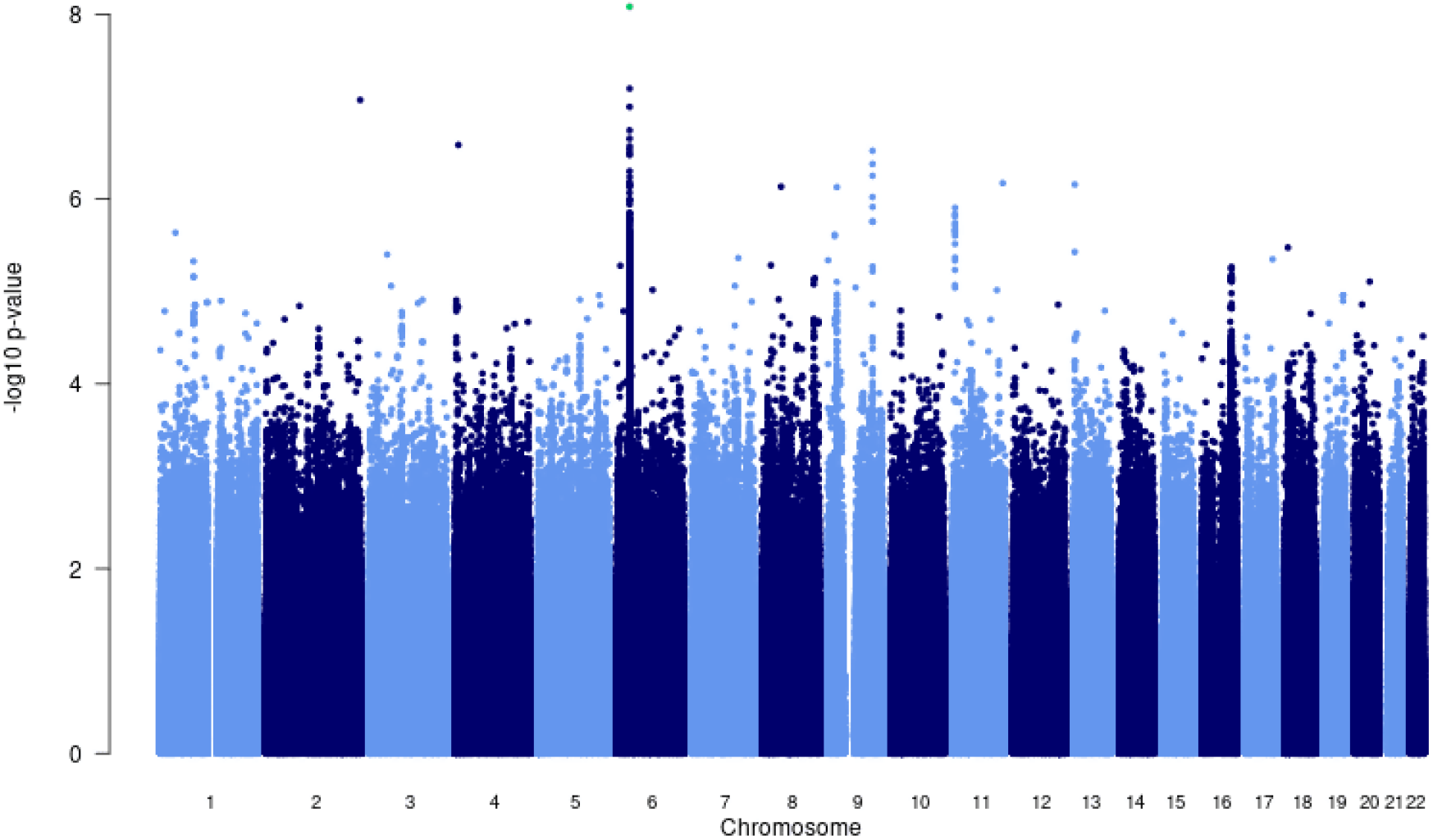
Manhattan plot of p-values (more than three studies) from the MR-Mega analysis of all 12 datasets with genomic control reveals one significant association in the *HLA-ll* region of chromosome 6 (rs28383206).

**Figure 2:**
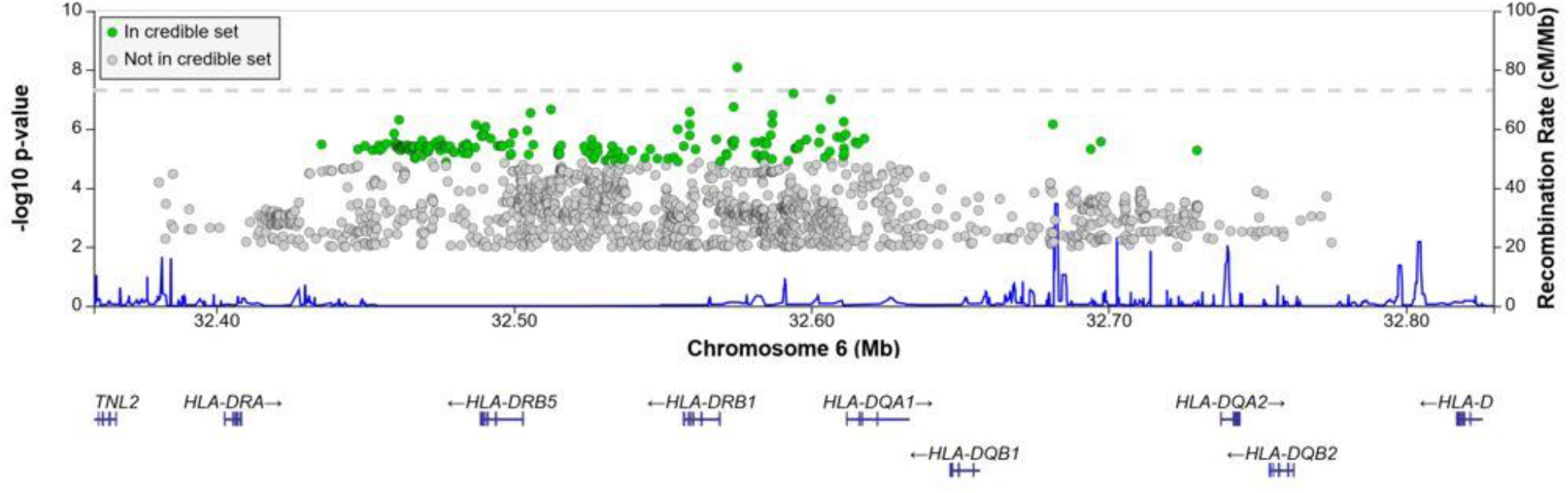
Regional association plot for the chromosome 6 *HLA-ll* rs28383206 association in the multi-ancestry analysis revealing a significant peak in the HLA-ll region. Image produced using the online LocusZoom database with LD mapping set to “all” and p-values > 0.01 removed^34^.

**Figure 3:**
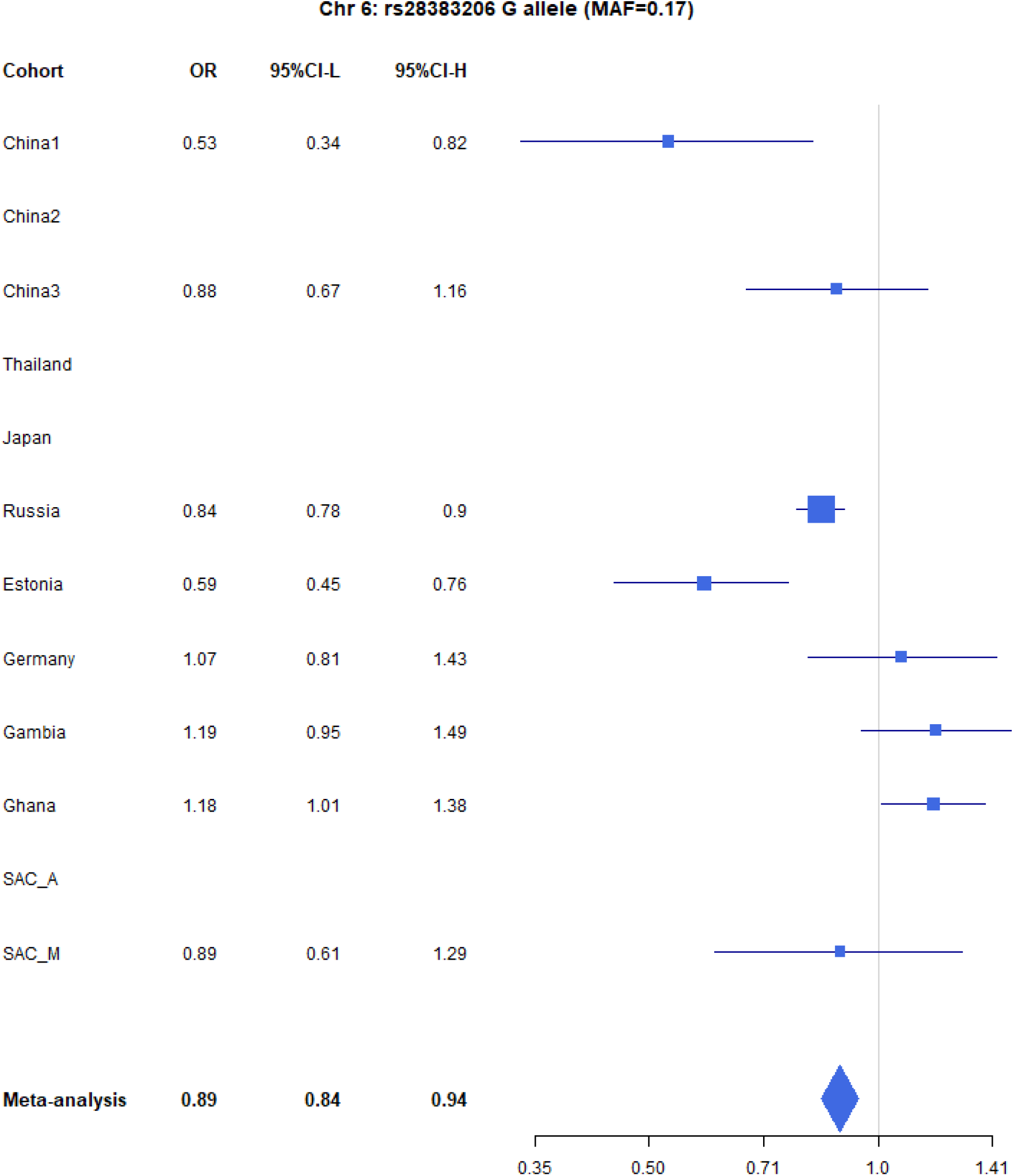
Forest plot (Odds ratio and 95% confidence interval) of the significant chromosome 6 association (rs28383206) for TB susceptibility in the multi-ancestry analysis, implemented using MR-Mega with GCC. Eight of the 12 studies included contained this variant. Studies that did not contain the variant are included in the plot but do not have results associated with them.

**Figure 4:**
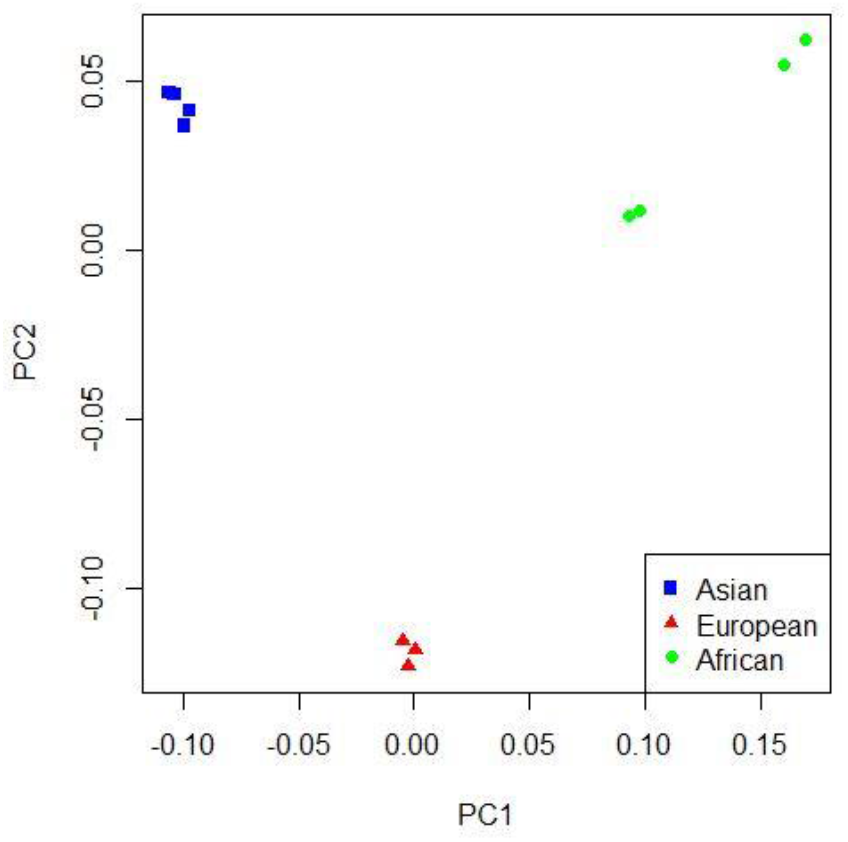
PCA plot of all 12 studies based on the MR-Mega mean pairwise genome-wide allele frequency differences

#### HLA-II

Given the strong association peak in the HLA-ll locus (Figure 2 and 3) we imputed HLA-ll alleles to fine-map this association. HLA alleles were imputed using the HIBAG R package which utilizes both genotyping array and population specific reference panels to obtain the most accurate imputations for each individual dataset. Association testing was then conducted using an additive genetic model for each individual dataset before meta-analyzing the results. After Bonferroni correction for all tested alleles across all HLA regions, multiple HLA-l and HLA-ll alleles demonstrated significant associations with disease for both the multi-ancestry and ancestry-specific analysis (supplementary data). The significant associations showed a strong consistency for direction of effects in all ancestry-specific meta-analysis, with the Asian and African having the least and most variation respectively, which is consistent with the higher genetic diversity within the African data. As expected, the multi-ancestry meta-analysis had more variation in effect sizes across all the input studies for the significant associations. However, some significant HLA class l and ll alleles presented with strong consistency in direction of effect across the input studies. Most notably, these associations included HLA-DQA1*03:01 and 03:03, also HLA-DQB1*03:02 (Figure S2), which have previously been reported in Russian and Icelandic populations^19^. Further, associations with high degree of overlap in effect direction were found in the HLA-DRB1, HLA-DPB1 and class 1 HLA-A, B and C genes, with the HLA class ll region having a greater number of associations with concordant direction of effect than the class l region. The high number of associated HLA alleles and consistency in effect direction for both the global and ancestry-specific analysis suggests a high degree of population-specific and globally shared TB susceptibility variants, which are exploitable by various *Mtb* strains across the globe to bring about disease. Sequencing efforts of global mycobacterial isolates find hyperconservation of HLA-II epitopes suggesting pathogen advantage achieved through HLA-II recognition and perhaps explaining risk associations of specific HLA variants^35^. The significant HLA associations overlap with the association peak observed in the multi-ancestry meta-analysis (Figure 4) but show more consistency in direction of effects between the input studies compared to the lead SNPs identified in the association peak. This suggests that the lead SNP in the meta-analysis is tagging an HLA allele, where the different LD patterns from the included ancestral populations result in the differences in effects sizes between populations at the lead SNP.

While the significant associations showed a high degree of consistent effect directions and similar alleles were under the topmost significant associations across all the ancestry-specific and multi-ancestry meta-analysis, there is still a large amount of variation between all the included studies, especially for variants that did not reach significance. Examining the HLA allele association statistics in the individual studies revealed that the top significant associations vary greatly across all included studies. This variation in significant associations is, in part, attributable to the observed variation in HLA allele frequencies across all the included studies (supplementary data). Differences in HLA allele frequencies were observed between different ancestries (Africa vs. Europe), indicating the vast variation within these loci. This variation could partly be due to unique infectious pressures that each geographical region faces and could also explain why different strains of *Mtb* are more, or less prevalent in different regions as they adapted to the HLA profile of the population within this region. Previous work has shown evidence of interaction between genetic variants of the host and specific strains of *Mtb* in Ghanaian and South African populations^7,8,36,37^. These interactions provide further evidence that *Mtb* may have undergone substantial genetic evolution, in concert with host migration and evolution of different populations^38,39^. Some studies suggest that HLA-II epitopes may have undergone regional mutations that modify HLA-II binding and we speculate that the heterogeneity observed in HLA-II associations between regions may, at least in part, be accounted for by different pressures exerted by varying stains of *Mtb* ^40^. However, part of this variation is likely due to differential tagging of underlying HLA alleles (or causal variants) by the SNPs available in the analysis. While imputation was done using genotyping array and population-specific reference panels, the available tagging SNPs will still vary by genotyping array, influencing the observed variation.

##### Impact of infection pressure on meta-regression

To further understand the heterogeneity across populations we attempted to account for variation in levels of prior exposure which could serve to mask host effects given that not all controls will have been exposed to *Mtb*. In low transmission settings, more susceptible but unexposed individuals would be included as controls, who had they been exposed to *Mtb* might have progressed to TB disease.

Overall, including each cohort’s estimated prevalence of prior exposure had a significant impact on the residual heterogeneity and association statistics of 5% of the variants included in the meta-analysis (419460/8355367), which at a significance level of p-value < 0.05 is what is to be expected purely by chance. Separating the results into bins according to p-values revealed that the bins where the covariate had the biggest impact was for p-values in the range of 1e^-3^ to 1e^-5^ (Figure S3), while significant and suggestive associations reported in this study did not show any significant changes in residual heterogeneity. While the proportion of variants significantly impacted is low and has the biggest impact on variants not significantly associated with TB susceptibility there was still an overall reduction in the chi-square value for the residual heterogeneity (mean chi-square value reduced by 10). This suggests that accounting for potential lifetime of infections does account for some of the observed residual heterogeneity, it is most likely not the main driving force for these residuals.

When considering the impact of force of infection, it is important to consider not only the proportion of controls ever exposed but also the impact of recurrent exposure. There is some evidence to suggest that genetic barriers to progression to TB may be overcome if the infectious dose is high^41^. Repeated exposure may be observed where TB prevalence is high, as in South Africa, and could contribute to the overall lower effects sizes observed in the GWAS enrolling SAC people. Inclusion of potential lifetime infections in meta-regression could help adjust for these effects and prove useful for not only TB, but meta-analysis of infectious diseases in general and should be further explored.

### Other suggestive loci that did not reach significance

There were four loci with suggestive associations and strong peaks on the Manhattan plot (Figure 1) that did not reach significance but should still be considered as potential variants of interest (Table S3). One chr9 peak (rs4576509) was intergenic (Figure S4) while the second (rs6477824) is located in the 5’-UTR region of the zinc finger protein 483 (*ZNF483*) gene (Figure S4), previously associated with age at menarche^42,43^. The chromosome 11 peak is located in the PPFIA binding protein 2 (*PPFIBP2*) gene (Figure S5), which plays a role in axon guidance and neuronal synapse development and has previously been implicated in cancer development^44,45^. The final peak, on chromosome 16 (Figure S6), is located in the craniofacial development protein 1 (*CFDP1*) gene region and involved in chromatin organization^46^. These genes have not been previously linked to TB susceptibility and a potential role is unclear and as a result further validation of these variants is needed before any conclusions on their impact to TB susceptibility can be drawn.

### Ancestry-specific meta-analysis

Concordance in direction of effects of the risk allele between the ancestry-specific meta-analyses were examined to determine if significant enrichment (above the expected 50%) exists at different p-value thresholds. Significant enrichment in concordance of direction of effect was only observed when using the European ancestry as reference compared to the African meta-analysis results for SNPs with p-values >0.001 and <0.01 (p-value = 0.0061, Table S4). The lack of enrichment between the ancestries suggests significant ancestry specific associations, which could be further compounded by differences in local infection pressures. Due to the lack of concordance and the separation of the ancestral populations in the PCA plot (Figure 4) ancestry-specific meta-analysis were done.

The PCA plot (Figure 4) for the 12 studies (based on mean pairwise genome-wide allele frequency differences calculated by MR-Mega) illustrates distinct separation between the three major population groups (Asia, Europe, Africa). The separation observed between the African studies (Gambia/Ghana and SAC) is due to the high level of admixture in the SAC population. The SAC population is a five-way admixed South African population with genetic contributions from Bantu-speaking African, KhoeSan, European, South and South East Asian populations, which explains the observed shift in the PCA plot^47^ (Figure 4).

QQ-plots for the ancestry-specific analysis show no significant inflation or deflation and after removing associations without any clear peaks on the Manhattan plots (associations driven by a single study) we found no significant associations for the ancestry-specific analysis. However, suggestive peaks that did not reach genome-wide significance were identified in the European and Asian ancestry-specific analyses (Figure S8 and S9). Potential causes for the lack of associations and suggestive peaks in the African analysis (Figure S10) are the increased genetic diversity within Africa and the inclusion of admixed samples (SAC), which could have resulted in a loss of power due to variation in allele frequencies. However, removing the admixed data and analyzing only the Gambian and Ghanaian datasets also did not produce any significant results. For the European analysis (Figure S8), suggestive peaks were identified on chromosome 6, 8, and 11 (Table S5), while the Asian (Figure S9) analysis identified suggestive peaks on chromosome 6 and 8 (Table S5).

The suggestive peaks on chromosome 6 and 11 in the European subgroup analysis overlap with the suggestive peaks of the multi-ancestry meta-analysis (Figure 1 and S7, Table S5), but the suggestive peak on chromosome 8 is unique to this population (Figure S8, Table S5). The peak is located in the ArfGAP with SH3 domain, ankyrin repeat and PH domain 1 (*ASAP1*) region, which encodes an ADP-ribosylation factor (ARF) GTPase-activating protein and is potentially involved in regulation of membrane trafficking and cytoskeleton remodeling^48^. Variants in *ASAP1* (rs4733781 and rs10956514) have previously been linked to TB susceptibility in a TB-GWAS analysis of the same Russian population included here^13^. While these ASAP1 variants were present in all twelve studies and had consistent direction of effects, they presented with a strong signal in the European ancestry-specific analysis only (African and Asian p-values all >=0.1). These differences in association were not driven by allele frequency differences as they are similar between the included study populations. A possible explanation for the association being observed only in the European meta-analysis could thus be a result of the increased sample size and power to detect associations in the European ancestry-specific analysis.

The suggestive peak on chromosome 8 in the Asian subgroup analysis lies in an intergenic region (Figure S9, Table S5) and the link to TB susceptibility is unclear. Finally, the suggestive region on chromosome 6 overlaps with the significant peak from the multi-ancestry analysis (Figure 1 and S9) and is located in the major histocompatibility complex, class II, DR beta 1 (*HLA-DRB1*), as discussed above (Figure S9, Table S5).

### Prior associations

To determine if associations from previously published TB-GWAS replicate in this meta-analysis we extracted all significant and suggestive associations from prior analyses and compared these to our multi-ancestry and ancestry-specific meta-analysis results^6,10–18,20,23–25,30,49^. In total, 44 SNPs and 36 genes were identified from the GWAS catalogue, of which 33 SNPs and all candidate genes were present in our data (supplementary data). We also extracted the association statistics for a further 90 previously identified candidate genes from our multi-ancestry and population specific meta-analysis results^3^.

Using a Bonferroni corrected p-value of 0.0015 for the number of SNPs tested (33) as the significance threshold for replication, two candidate SNPs (rs4733781: p-value = 3.22e-05; rs10956514: p-value = 0.000118; supplementary data) replicated in the multi-ancestry meta-analysis, both located in the *ASAP1* gene region^13,50,51^. Given the overlap between the multi-ancestry meta-analysis and the GWAS in which these SNPs were discovered, it is not surprising that they also present with suggestive associations in this study. As discussed above however, these variants did not show a strong signal in any ancestry-specific analysis except for Asia, and the combined multi-ancestry analysis reveals that the p-values are lower when compared to the European ancestry-specific analysis. As the Russian cohort from Curtis *et al*. (2015) is included in this analysis, it is not surprising that these variants replicate in the European ancestry-specific analysis. The lack of signal for these variants in the Asian and African cohorts and reduced p-value in the multi-ancestry analysis means that the European data drove the association of these *ASAP1* variants.

For the Asian ancestry-specific analysis, the replicated variant was rs41553512, located in the *HLA-DRB5* gene (p-value = 3.53E-05). *HLA-DRB5* is located within the HLA-ll region identified in the multi-ancestry meta-analysis (Figure 2) and was previously identified by Qi *et al*. (2017) in a Han Chinese population. The African ancestry-specific analysis did not replicate previous associations, with the lowest p-value at rs6786408 in the *FOXP1* gene (p-value = 0.023). While this variant was previously identified in a North African cohort, the fact that it does not replicate here could be because of the genetic diversity within Africa and specifically the variability introduced by the five-way admixed South African population.

## Discussion

This first ever large-scale, multi-ethnic meta-analysis of genetic susceptibility to TB, involving 14153 cases and 19536 controls, identified one risk locus achieving genome-wide significance and further investigation of this region revealed significant HLA epitopes. This association is noteworthy as HLA has consistently been associated with TB susceptibility across multiple populations, although not involving similar alleles^28–30^.

While the significant association identified in this multi-ancestry analysis does present with a high level of heterogeneity (Figure 3) in genetic effects between the source populations, further analysis of the HLA locus identified multiple, significantly associated, HLA epitopes with consistent direction of effects and less heterogeneity. The consistency of effects for these HLA epitopes suggests a degree of globally shared TB susceptibility that should be explored further. However, while consistency in direction of effects was observed for some HLA epitopes, many still presented with high levels of heterogeneity and when comparing HLA epitope frequencies between the input studies, it is evident that the dominant epitopes vary between the included studies.

*Mtb* was a human pathogen prior to human migration out of Africa, which supports the finding of a globally associated TB susceptibility variant, as identified here^38^. Subsequent host pathogen co-evolution and the emergence of dominant lineages in different populations^52^ could have driven the underlying heterogeneity observed between the studies by selecting for different HLA epitopes^8,39,40^.

The p-values of residual heterogeneity in genetic effects between the studies in the multi-ancestry meta-analysis show no significant inflation between the studies suggesting that differences in study design (phenotype definition, infection pressure, Mtb strain) are not a major contributor to the lack of significant associations. However, the ancestry-correlated heterogeneity p-values are generally lower than the residual heterogeneity, suggesting that genetic ancestry has a stronger impact on the differences in effects sizes between the studies. For the ancestry-specific analysis, fewer studies result in there being less input heterogeneity to account for, but the reduced sample size was not sufficient to detect any ancestry-specific genome-wide associations. This is particularly evident for the African ancestry-specific meta-analysis where the large degree of heterogeneity, which could be a result of the high genetic diversity within Africa, resulted in no observable suggestive association peaks^53,54^. Furthermore, the suggestive associations reported in this study should be interpreted with care and further validation is required before any conclusions can be drawn on the impact that they could have on TB susceptibility.

Polygenic heritability estimates revealed genetic contributions to TB susceptibility for all studies, but the level of this contribution varied greatly (5-36%), suggesting that other factors are contributing to both the lack of significant associations detected in this meta-analysis and the variation observed for the polygenic heritability estimates. These factors likely include environmental, socioeconomic, and varying levels of infection pressures, as well as genetic ancestry specific effects between the included study populations. An individual from South Africa will face a much higher force of infection than individuals in Europe, and making the assumption that environmental circumstances are equal will significantly skew these crude heritability estimates^26^. This argument is sustained by the fact that increasing disease prevalence (higher infection pressure) increased the level of genetic contribution to TB susceptibility up to a certain point, after which further increasing the infection pressure will not further impact genetic susceptibility.

To determine the impact that force of infection has on the level of genetic contribution to TB susceptibility we modeled values for proportion of people ever infected with *Mtb* to include in the multi-ancestry meta-analysis and correct for the different force of infection faced by individuals in each country. Inclusion of this covariate however only resulted in a significant difference for 5% of the analyzed variants, what is to be expected based on chance alone and as such we cannot conclude that a significant portion of the observed residual heterogeneity is explained by this. Limited meta-data forced us to make a number of assumptions about the ages of study participants and the dates on which they were enrolled. With more precise meta-data, or *Mtb* infection test results in controls, the potential impact of lifetime infection could be better quantified and may contribute to elucidating genetic TB susceptibility. Multi-ancestry meta-analysis of other infectious diseases could also potentially benefit from inclusion of force of infection covariates. It would also be important to determine whether there is a level of exposure beyond which host genetic barriers to infection are overcome^55^.

While a significant association was identified in this multi-ancestry meta-analysis, the number of associations is small when compared to other meta-analyses of similar sizes. Factors contributing to this include the difficulty of analyzing multi-ancestry data, the outdated arrays and lack of suitable reference panels for included study populations, and heterogeneity in case and control definitions between the studies. The issue of heterogeneity in definitions is especially pronounced for this study as it included unpublished data with limited information, which does not indicate how cases were confirmed and controls were collected. The complexity of TB and generally small genetic effects suggest that larger sample sizes or alternative methods of investigation are needed. Utilizing GWAS arrays that better capture diverse populations in combination with imputation making use of larger and more diverse reference panels would allow for larger and more consistent datasets for future meta-analysis. Remapping of specific areas of interest such as the *HLA, ASAP1* or *TLR* using long read sequencing would be invaluable. Increased amounts of genetic data will also allow for more accurate TB heritability analysis and permit analysis of polygenic risk scores and exploration of host pathogen interactions.

In conclusion, this first large-scale multi-ancestry TB GWAS meta-analysis revealed significant associations and shared genetic TB susceptibility architecture across multiple populations from different genetic backgrounds. The analysis shows the value of collaboration and data sharing to solve difficult problems and elucidate what determines susceptibility to complex diseases such as TB. We hope that this publication will encourage others to make their data available for future large scale meta-analysis.

## Methods

### Data

This analysis includes 12 of the 17 published (and un-published, Table 1 and S1) GWAS studies of TB (with HIV negative cohorts) prior to 2022^10–17,49^. It excludes data from Iceland and Vietnam^18,30^, as they declined to share data. It excludes data from China, Korea, Peru and Japan^6,20,21,23,30^, as data sharing agreements could not be finalized in time for this analysis. The Indonesian data was not suitable for reliable imputation and the Moroccan data was family-based and thus also not suitable for this meta-analysis^24,25^. Finally, genotyped TB cases and controls are also available in the UK Biobank, but this data was not included in this analysis as genetic association studies on such highly selected datasets need to be undertaken with caution and to not bias results were excluded for this analysis^56^.

Included individuals were genotyped on a variety of genotyping arrays (Table 1 and S1), and raw genotyping data was available for eight datasets and for the remainder association testing summary statistics were obtained to use in the meta-analysis (Table 1 and S1). Quality control (QC) and imputation of the data with raw genotyping information available (Table 1 and S1) was done using Plink (v1.9), followed by pre-phasing using SHAPEIT and Impute2 with the 1000 genomes phase 3 reference panel^57–60^. QC and imputation was done as described previously^10,61^, briefly we used a minor allele filter of 0.025 and an individual and SNP missingness filter of 0.1. Hardy–Weinberg equilibrium threshold was set at a Bonferroni corrected p-value according to the number of SNPs testes (0.05/number of SNPs) and samples where sex could not be determined from genotyping were also removed. Imputed data was filtered at a quality score of 0.3, prior to individual and genotype filtration steps. Prior to QC and imputation, allele orientation was corrected using Genotype Harmoniser version 1.4.15 and the genome build of all datasets was checked for consistency (GRCh37) and updated if necessary using the liftOver software from the UCSC genome browser^62,63^. The four datasets with only summary statistics available (Table 1 and S1) were imputed and QC’d during the original investigations, but the marker names and allele orientation were checked for concordance between the summary statistics and the rest of the consortium’s imputed data.

### Polygenic heritability analysis

To assess the level of genetic contribution to TB susceptibility we estimated Polygenic heritability on the individual studies for which raw genotyping data was available (Table 1 and S1). Polygenic heritability estimates were calculated using GCTA (v1.93.2), a genomic risk prediction tool^64^. The genetic relationship matrix was calculated for each autosomal chromosome prior to being merged and filtered by removing cryptic relatedness (--grm-cutoff 0.025) and assuming that the causal loci have similar distribution of allele frequencies as the genotyped SNPs (--grm-adj 0). Principal components were then calculated (--pca 20) to include as covariates prior to estimating heritability. The average heritability estimate was calculated by taking the mean of all estimates and the confidence intervals were estimated based on the standard error across all studies and the number of studies included.

### Meta-analysis

All variants with minor allele frequency (MAF) >1% and polymorphic in at least three studies (from at least two different ancestries) were included in the primary analysis. For the GWAS summary statistics of each dataset variants with infinite confidence intervals were removed prior to the meta-analysis. A multi-ancestry meta-analysis plus separate ancestry-specific analyses for Africa, Asia and Europe were performed. MR-MEGA (Meta-Regression of Multi-Ethnic Genetic Association, v0.20), a meta-analysis tool that maximizes power and enhances fine-mapping when combining data across different ethnicities, was used for the multi-ancestry meta-analysis^65^. To account for the expected heterogeneity in allelic effects between populations, MR-MEGA implements a multi-ancestry meta-regression that includes covariates to represent genetic ancestry, obtained from multi-dimensional scaling of mean pairwise genome-wide allele frequency differences. Genomic control correction (GCC) was implemented during the MR-MEGA analysis for the individual input data (if lambda was >1.05) and output statistics, and the first two principal components (PCs), calculated from the genome-wide allele frequency differences, were included as covariates in the regression. QQ-plots of p-values and associated lambda values were used to assess the quality of results prior to downstream investigation.

For the ancestry-specific analyses, the studies were grouped by the major ancestral groups (Table 1 and S1) and all variants with a MAF of >1% that were observed in at least 2 studies were included in the meta-analysis. We performed traditional fixed effects meta-analyses in GWAMA (v2.2.2), implementing GCC, and assessed the results using QQ-plots^66^. The genome-wide significance threshold for all association testing was set at p-value <= 5×10^−8 67^.

### HLA imputation

To fine-map *HLA* alleles over the *HLA* locus we imputed *HLA class l and ll* variants for all studies for which raw data was available (Table 1 and S1). *HLA* imputation for the *HLA class l* regions *A, B* and *C* as well as the *HLA class ll* regions *DPB1, DRB1, DQB1* and *DQA1* was done using the R package HIBAG (version 1.5), implemented in the R free software environment (version 4.0.5) using the predict() command for imputation^68–70^.

The reference datasets for HLA imputation are both genotyping panel and population specific and HIBAG has a database of reference data for many genotyping arrays. Each reference panel is also available for either Asian, European or African populations or a mixture of the three (https://hibag.s3.amazonaws.com/hlares_index.html#estimates). For each dataset included for imputation the reference panel chosen was the same as the genotyping array used for the data and the reference population was chosen to match the data as closely as possible. Asian and European reference panels were used for the Asian and European populations and African references were used for the Gambia and Ghana datasets, while mixed datasets were implemented for the admixed SAC population.

Following imputation, the HIBAG package (hlaAssocTest) command was used to implement an additive association test for the HLA alleles across the different regions. Association testing results for the eight included studies were then combined with a restricted maximum-likelihood estimator model meta-analysis using the R package metafor (version 2.4-0). Ancestry-specific meta-analysis grouped according to the major population groups (Table 1 and S1) were also done using the same method. Meta-analysis models were chosen based on heterogeneity in the data.

For a heterogeneity result of *p*-value > 0.1 the fixed effects (FE) model (inverse-variance method) was implemented and for *p*-value ≤ 0.1 the random effects (RE) model (restricted maximum likelihood estimator) was used to calculate pooled OR and CI values^71^.

#### Estimation of infection pressure

To generate a covariate capturing the likely cumulative exposure to *Mtb* for included controls, the results of Houben and Dodd (2016) were adapted to produce a distance matrix to feed into the meta-analysis. A sample of 200 estimated histories of the annual risk of TB infection in each country was used to calculate the expected fraction of control participants ever infected with *Mtb*, assuming that controls were uniformly aged between 35-44 years in 2010. This was done by including estimates for the potential lifetime infections for each source population as a covariate in the MR-MEGA multi-ancestry meta regression. To determine the impact of the covariate a chi-square difference test was implemented, on a SNP-SNP basis, on the residual and association testing statistics of two meta-analysis output statistics, one including and the other excluding the potential lifetime infections covariate^72^. The aim was to determine if inclusion of potential lifetime infections in the regression explained some of the residual heterogeneity.

#### Concordance of direction of effect

To determine the degree to which direction of effect is shared for SNPs between the ancestry specific meta-analysis we followed the methodology of Mahajan *et. al*.^73^. First identified all variants present in all 12 included datasets. Among these SNPs we then identified an independent subset of variants in the European ancestry-specific meta-analysis showing nominal evidence of association (p-value <= 0.001) and separated by at least 500kb. The identified SNPs were then extracted from the Asian and African ancestry-specific meta-analysis results to calculate the number of SNPs that had the same direction of effect as in the European analysis. To determine if significant excess in concordance of effect direction was present a one-sided binomial test was implemented with the expected concordance set at 50%. This analysis was then repeated for other p-value thresholds (0.001 < P ≤ 0.01; 0.01 < P ≤ 0.5; and 0.5 < P ≤ 1), and also using the African and Asian meta-analysis results as reference.

## Data Availability

Summary statistics of all analysis will be made available on the NHGRI-EBI Catalog of human genome-wide association studies database upon official publication of the manuscript, but the summary statistics and original data can be requested through the corresponding authors.

## Conflict of interest

The authors declare no conflict of interest.

## Ethics statement

A research collaboration agreement was signed by all contributors. Ethics approval for the meta-analysis presented here was granted by the Health Research Ethics Committee of Stellenbosch University (project registration number S17/01/013). In addition, all institutions involved in the ITHGC have ethics approval for their respective studies:

China 1 and 2: The study protocol was approved by the Ethics Committee of the Beijing Chest Hospital, the 309 Hospital of the PLA, Shijiazhuang Fifth Hospital, the China PLA General Hospital, the Tongliao TB institute and the Center for Diseases Control and Prevention in Jalainuoer.

China 3: Ethics approval was granted by the Ethics Committees of the Beijing Children’s Hospital, the Beijing Geriatric Hospital, the Tuberculosis Hospital in Shaanxi Province, the Beijing Institute of Genomics, Chinese Academy of Sciences and the Center for Disease Control and Prevention of Jiangsu Province.

Thailand: Ethics approval was granted by the Ethics Review Committee of the Ministry of Public Health in Thailand.

Japan: Ethics approval was granted by the Institutional Review Board of the Center for Genomic Medicine, RIKEN

Russia: Blood samples from all participants were collected and studied with written informed consent according to the Declaration of Helsinki and with approvals from the local ethics committees in Russia (St. Petersburg and Samara) and the UK (Human Biological Resource Ethics Committee of the University of Cambridge and the National Research Ethics Service, Cambridgeshire 1 REC, 10/H0304/71).

Estonia: The Estonian Bioethics and Human Research Council (EBIN) approved the Estonian Genome Center study reported in this manuscript.

Germany: The study protocol was approved by the ethics committee (EC) of the University of Luebeck, Germany (reference 07–125), and was adopted by other ethics committees covering all 18 participating centres (EC of the medical faculty of the University of Goettingen; EC of the Medical Council of Hessen, Frankfurt /Main; EC of the Medical Council Hamburg; EC of the Medical Council Lower Saxony, Hannover; EC of the Medical Faculty Carl Gustav Carus, Technical University of Dresden; EC of the Medical Council Berlin; EC of the Medical Council Bavaria, Munich; EC of the Medical Faculty, Friedrich-Alexander-University Erlangen-Nuremberg; EC of the Medical Faculty of the University of Regensburg; EC of the University of Witten/ Herdecke) Gambia: Ethics approval was granted by the Medical Research Council (MRC) and the Gambian government joint ethical committee.

Ghana: Ethics approval was granted by the Committee on Human Research, Publications and Ethics, School of Medical Sciences, Kwame Nkrumah University of Science and Technology, Kumasi, Ghana, and the Ethics Committee of the Ghana Health Service, Accra, Ghana.

SAC A and SAC M: Ethics approval was granted by the Health Research Ethics Committee of Stellenbosch University (project registration numbers S17/01/013, NO6/07/132 and 95/072).

## Acknowledgement

Computation used the Oxford Biomedical Research Computing (BMRC) facility, a joint development between the Wellcome Centre for Human Genetics and the Big Data Institute supported by Health Data Research UK and the NIHR Oxford Biomedical Research Centre. Financial support was provided by the Wellcome Trust Core Award Grant Number 203141/Z/16/Z. The views expressed are those of the author(s) and not necessarily those of the NHS, the NIHR or the Department of Health and Social Care. This work was partly supported by a Grant in-Aid for Scientific Research (B), (KAKENHI 21406006) from Japan Society for the Promotion of Science (JSPS).

The clinical information and samples in Thailand, in this part was supported by JSPS KAKENHI 17256005 and later by research grant from Ministry of Health, Labor and Welfare (MHLW) H21-aids-12. We would like to thank all the subjects and the members of the Rotary Club of Osaka-Midosuji District 2660 Rotary International in Japan who donated their DNA for this work. We thank all members of BioBank Japan, Institute of Medical Science, The University of Tokyo and of RIKEN Center for Genomic Medicine for their contribution to the completion of our study. This work was conducted as a part of the BioBank Japan Project that was supported by the Ministry of Education, Culture, Sports, Science and Technology of the Japanese government. As for Thai samples, we thank all of the staffs and collaborators of the TB/HIV Research Project, Thailand, a research project between the Research Institute of Tuberculosis, the Japan Anti-tuberculosis Association and the Thai Ministry of Public Health for collecting clinical data and DNA samples. We thank the German Consortium “TB or not TB Network” (https://www.tbornottb.de/) which was responsible for collecting the German TB samples. We acknowledge the support of the DSI-NRF Centre of Excellence for Biomedical Tuberculosis Research, South African Medical Research Council Centre for Tuberculosis Research, Division of Molecular Biology and Human Genetics, Faculty of Medicine and Health Sciences, Stellenbosch University, Cape Town, South Africa.

## Funding

JJG is funded by an NIHR Academic Clinical Lectureship. A.P.M. acknowledges support from Versus Arthritis (grant reference 21754). PJD was supported by a fellowship from the UK Medical Research Council (MR/P022081/1); this UK funded award is part of the EDCTP2 program supported by the European Union. ME was supported by an NHMRC fellowship (552496). The research was supported by the NHMRC grant 1025166. AvL and RvC are supported by the National Institute of Allergy and Infectious Diseases at NIH [R01 AI136921]. TAY is an NIHR Clinical Lecturer supported by the National Institute for Health Research. TP acknowledges funding from the National Institute for Health Research (CL-2020-21-001) and the Wellcome Trust (222098/Z/20/Z). The views expressed in this publication are those of the author(s) and not necessarily those of the NHS, the National Institute for Health Research or the Department of Health and Social Care. AM and RM are funded by the EU project no. 2014-2020.4.01.15-0012 “Gentransmed”. BA is supported by the ‘Scientific Programme Indonesia Netherlands’ (SPIN) under the Royal Academy of Arts and Sciences (KNAW), the Netherlands.

## The International Tuberculosis Host Genetics Consortium

### Central analysis team

Haiko Schurz^1^*, Vivek Naranbhai^2,3,4,5,6^*^#^, Tom A. Yates^7^, James J. Gilchrist^2,8^, Tom Parks^2,9^, Peter J. Dodd^10^, Marlo Möller^1^, Eileen G Hoal^1^, Andrew P. Morris^11^, Adrian V.S. Hill^2,12#^

### Collaborators

Reinout van Crevel^13,14^, Arjan van Laarhoven^13^, Tom H.M. Ottenhoff^15^, Andres Metspalu^16^, Reedik Magi^16^, Christian G. Meyer^17^, Magda Ellis^18^, Thorsten Thye^19^, Surakameth Mahasirimongkol^20^, Ekawat Pasomsub^21^, Katsushi Tokunaga^22^, Yosuke Omae^22^, Hideki Yanai^23^, Taisei Mushiroda^24^, Michiaki Kubo^24^, Atsushi Takahashi^25,26^, Yoichiro Kamatani^25^, Bachti Alisjahbana^27^, Wei Liu^28,29^, A-dong Sheng^30,31,32,33^, Yurong Yang^34^

### Affiliations

1. DSI-NRF Centre of Excellence for Biomedical Tuberculosis Research, South African Medical Research Council Centre for Tuberculosis Research, Division of Molecular Biology and Human Genetics, Faculty of Medicine and Health Sciences, Stellenbosch University, Cape Town, South Africa
2. Wellcome Trust Centre for Human Genetics, Oxford, UK
3. Massachusetts General Hospital, Boston, USA
4. Dana-Farber Cancer Institute, Boston, USA
5. Centre for the AIDS Programme of Research in South Africa, Durban, South Africa
6. Harvard Medical School, Boston, USA
7. Division of Infection and Immunity, Faculty of Medical Sciences, University College London, London, UK
8. Department of Paediatrics, University of Oxford, UK.
9. Department of Infectious Diseases Imperial College London, London, UK
10. School of Health and Related Research, University of Sheffield, Sheffield, UK
11. Centre for Genetics and Genomics Versus Arthritis, Centre for Musculoskeletal Research, The University of Manchester, Manchester, UK.
12. Jenner Institute, University of Oxford, UK.
13. Department of Internal Medicine and Radboud Center for Infectious Diseases, Radboud University Medical Center, Nijmegen, the Netherlands
14. Centre for Tropical Medicine and Global Health, Nuffield Department of Medicine, University of Oxford, Oxford, UK
15. Head Lab Dept of Infectious Diseases; Head Group Immunology and Immunogenetics of Bacterial Infectious Diseases Leiden University Medical Center, PO Box 9600, 2300RC Leiden, the Netherlands
16. Estonian Genome Center, Institute of Genomics, University of Tartu, Estonia.
17. Institute of Tropical Medicine, Eberhard-Karls University Tübingen, Germany and Faculty of Medicine, Duy Tan University, Da Nang, Vietnam
18. Tuberculosis Research Group, Centenary Institute, Sydney, Australia.
19. Department of Infectious Disease Epidemiology, Bernhard Nocht Institute for Tropical Medicine Hamburg, 20359 Hamburg, Germany.
20. Department of Medical Sciences, Ministry of Public Health, Nonthaburi, 11000, Thailand
21. Virology Laboratory, Department of Pathology, Faculty of Medicine, Ramathibodi Hospital, Mahidol University, Bangkok 10400, Thailand
22. Genome Medical Science Project, National Center for Global Health and Medicine, Tokyo, 162-8655, Japan.
23. Fukujuji Hospital and Research Institute of Tuberculosis, Japan Anti-Tuberculosis Association, Kiyose, 204-0022, Japan.
24. RIKEN Center for Integrative Medical Sciences, Yokohama, Kanagawa, 230-0045, Japan
25. Laboratory for Statistical Analysis, RIKEN Center for Integrative Medical Sciences, Yokohama, Kanagawa, 230-0045, Japan
26. Department of Genomic Medicine, Research Institute, National Cerebral and Cardiovascular Center, Suita, Osaka, 564-8565, Japan
27. Faculty of Medicine, Universitas Padjdjaran - Hasan Sadikin Hospital, Bandung Indonesia.
28. Department of Plastic and reconstructive surgery, shanghai Key laboratory of Tissue engineering, shanghai Ninth People’s hospital, shanghai Jiao Tong University – school of Medicine, shanghai, People’s republic of China
29. National Tissue engineering center of China, shanghai, People’s republic of China
30. National Clinical Research Center for Respiratory Diseases, National Key Discipline of Pediatrics, Capital Medical University, No. 56 Nanlishi Road, Xicheng District, Beijing, 100045, China.
31. Key Laboratory of Major Diseases in Children, Ministry of Education, Beijing Children’s Hospital, National Center for Children’s Health, Capital Medical University, No. 56 Nanlishi Road, Xicheng District, Beijing, 100045, China.
32. Beijing Key Laboratory of Pediatric Respiratory Infection Diseases, Beijing Pediatric Research Institute, No. 56 Nanlishi Road, Xicheng District, Beijing, 100045, China.
33. Children’s Hospital Affiliated to Zhengzhou University, Henan Children’s Hospital, Zhengzhou Children’s Hospital, Zhengzhou, China.
34. Ningxia Medical University, Ningxia Hui Autonomous Region, the People’s Republic of China

## Notes

### Competing Interest Statement

The authors have declared no competing interest.

### Author Declarations

A research collaboration agreement was signed by all contributors. Ethics approval for the meta analysis presented here was granted by the Health Research Ethics Committee of Stellenbosch University (project registration number S17/01/013). In addition, all institutions involved in the ITHGC have ethics approval for their respective studies: China 1 and 2: The study protocol was approved by the Ethics Committee of the Beijing Chest Hospital, the 309 Hospital of the PLA, Shijiazhuang Fifth Hospital, the China PLA General Hospital, the Tongliao TB institute and the Center for Diseases Control and Prevention in Jalainuoer. China 3: Ethics approval was granted by the Ethics Committees of the Beijing Childrens Hospital, the Beijing Geriatric Hospital, the Tuberculosis Hospital in Shaanxi Province, the Beijing Institute of Genomics, Chinese Academy of Sciences and the Center for Disease Control and Prevention of Jiangsu Province. Thailand: Ethics approval was granted by the Ethics Review Committee of the Ministry of Public Health in Thailand. Japan: Ethics approval was granted by the Institutional Review Board of the Center for Genomic Medicine, RIKEN Russia: Blood samples from all participants were collected and studied with written informed consent according to the Declaration of Helsinki and with approvals from the local ethics committees in Russia (St. Petersburg and Samara) and the UK (Human Biological Resource Ethics Committee of the University of Cambridge and the National Research Ethics Service, Cambridgeshire 1 REC, 10/H0304/71). Estonia: The Estonian Bioethics and Human Research Council (EBIN) approved the Estonian Genome Center study reported in this manuscript. Germany: The study protocol was approved by the ethics committee (EC) of the University of Luebeck, Germany (reference 07/125), and was adopted by other ethics committees covering all 18 participating centres (EC of the medical faculty of the University of Goettingen; EC of the Medical Council of Hessen, Frankfurt /Main; EC of the Medical Council Hamburg; EC of the Medical Council Lower Saxony, Hannover; EC of the Medical Faculty Carl Gustav Carus, Technical University of Dresden; EC of the Medical Council Berlin; EC of the Medical Council Bavaria, Munich; EC of the Medical Faculty, Friedrich Alexander University Erlangen Nuremberg; EC of the Medical Faculty of the University of Regensburg; EC of the University of Witten/ Herdecke) Gambia: Ethics approval was granted by the Medical Research Council (MRC) and the Gambian government joint ethical committee. Ghana: Ethics approval was granted by the Committee on Human Research, Publications and Ethics, School of Medical Sciences, Kwame Nkrumah University of Science and Technology, Kumasi, Ghana, and the Ethics Committee of the Ghana Health Service, Accra, Ghana. SAC A and SAC M: Ethics approval was granted by the Health Research Ethics Committee of Stellenbosch University (project registration numbers S17/01/013, NO6/07/132 and 95/072).

### Summary of Updates

The revised document has changes to the funding sections and for some of the co-authors the names have been updated (added middle name initial), the format of the author list has also been updated and a consortium heading has been added to list all individuals in the authors of the consortium.

## References

1. Houben, R. M. G. J. & Dodd, P. J. The Global Burden of Latent Tuberculosis Infection: A Re-estimation Using Mathematical Modelling. PLoS Med. 13, e1002152 (2016).

2. WHO Global tuberculosis report 2020. https://www.who.int/publications/i/item/9789240013131.

3. Naranbhai, V. The Role of Host Genetics (and Genomics) in Tuberculosis. Microbiol. Spectr. 4, (2016).

4. Correa-Macedo, W., Cambri, G. & Schurr, E. The Interplay of Human and Mycobacterium Tuberculosis Genomic Variability. Front. Genet. 10, 865 (2019).

5. Daya, M. et al. Using multi-way admixture mapping to elucidate TB susceptibility in the South African Coloured population. BMC Genomics 15, 1021 (2014).

6. Luo, Y. et al. Early progression to active tuberculosis is a highly heritable trait driven by 3q23 in Peruvians. Nat. Commun. 10, (2019).

7. Möller, M. & Kinnear, C. J. Human global and population-specific genetic susceptibility to Mycobacterium tuberculosis infection and disease. Curr. Opin. Pulm. Med. 26, 302–310 (2020).

8. Müller, S. J. et al. A multi-phenotype genome-wide association study of clades causing tuberculosis in a Ghanaian- and South African cohort. Genomics 113, 1802–1815 (2021).

9. Omae, Y. et al. Pathogen lineage-based genome-wide association study identified CD53 as susceptible locus in tuberculosis. J. Hum. Genet. 62, 1015–1022 (2017).

10. Schurz, H. et al. A Sex-Stratified Genome-Wide Association Study of Tuberculosis Using a Multi-Ethnic Genotyping Array. Front. Genet. 9, (2019).

11. Chimusa, E. R. et al. Genome-wide association study of ancestry-specific TB risk in the South African Coloured population. Hum. Mol. Genet. 23, 796–809 (2014).

12. Consortium, T. W. T. C. C. Genome-wide association study of 14,000 cases of seven common diseases and 3,000 shared controls. Nature 447, 661 (2007).

13. Curtis, J. et al. Susceptibility to tuberculosis is associated with variants in the ASAP1 gene encoding a regulator of dendritic cell migration. Nat. Genet. 47, 523–527 (2015).

14. Mahasirimongkol, S. et al. Genome-wide association studies of tuberculosis in Asians identify distinct at-risk locus for young tuberculosis. J. Hum. Genet. 57, 363–367 (2012).

15. Qi, H. et al. Discovery of susceptibility loci associated with tuberculosis in Han Chinese. Hum. Mol. Genet. 26, 4752–4763 (2017).

16. Thye, T. et al. Genome-wide association analyses identifies a susceptibility locus for tuberculosis on chromosome 18q11.2. Nat. Genet. 42, 739–741 (2010).

17. Thye, T. et al. Common variants at 11p13 are associated with susceptibility to tuberculosis. Nat. Genet. 44, 257–259 (2012).

18. Quistrebert, J. et al. Genome-wide association study of resistance to Mycobacterium tuberculosis infection identifies a locus at 10q26.2 in three distinct populations. PLOS Genet. 17, e1009392 (2021).

19. Sveinbjornsson, G. et al. HLA class II sequence variants influence tuberculosis risk in populations of European ancestry. Nat. Genet. 48, 318–322 (2016).

20. Hong, E. P., Go, M. J., Kim, H.-L. & Park, J. W. Risk prediction of pulmonary tuberculosis using genetic and conventional risk factors in adult Korean population. PLOS ONE 12, e0174642 (2017).

21. Li, M. et al. A next generation sequencing combined genome-wide association study identifies novel tuberculosis susceptibility loci in Chinese population. Genomics 113, 2377– 2384 (2021).

22. Luo, Y. et al. Early progression to active tuberculosis is a highly heritable trait driven by 3q23 in Peruvians. Nat. Commun. 10, 3765 (2019).

23. Zheng, R. et al. Genome-wide association study identifies two risk loci for tuberculosis in Han Chinese. Nat. Commun. 9, 4072 (2018).

24. Grant, A. V. et al. A genome-wide association study of pulmonary tuberculosis in Morocco. Hum. Genet. 135, 299–307 (2016).

25. Png, E. et al. A genome wide association study of pulmonary tuberculosis susceptibility in Indonesians. BMC Med. Genet. 13, 5 (2012).

26. Pearce, N. Epidemiology in a changing world: variation, causation and ubiquitous risk factors. Int. J. Epidemiol. 40, 503–512 (2011).

27. Thu, K. S. et al. Association of polymorphisms of the transporter associated with antigen processing (TAP2) gene with pulmonary tuberculosis in an elderly Japanese population. APMIS Acta Pathol. Microbiol. Immunol. Scand. 124, 675–680 (2016).

28. Kinnear, C., Hoal, E. G., Schurz, H., van Helden, P. D. & Möller, M. The role of human host genetics in tuberculosis resistance. Expert Rev. Respir. Med. 11, 721–737 (2017).

29. Stein, C. M. et al. Genomics of human pulmonary tuberculosis: from genes to pathways. Curr. Genet. Med. Rep. 5, 149–166 (2017).

30. Sveinbjornsson, G. et al. HLA class II sequence variants influence tuberculosis risk in populations of European ancestry. Nat. Genet. 48, 318–322 (2016).

31. Zhang, M. et al. Associations of genetic variants at TAP1 and TAP2 with pulmonary tuberculosis risk among the Chinese population. Epidemiol. Infect. 149, e79 (2021).

32. Kelly, A. & Trowsdale, J. Genetics of antigen processing and presentation. Immunogenetics 71, 161–170 (2019).

33. Shiina, T., Hosomichi, K., Inoko, H. & Kulski, J. K. The HLA genomic loci map: expression, interaction, diversity and disease. J. Hum. Genet. 54, 15–39 (2009).

34. Boughton, A. P. et al. LocusZoom.js: interactive and embeddable visualization of genetic association study results. Bioinformatics 37, 3017–3018 (2021).

35. Comas, I. et al. Human T cell epitopes of Mycobacterium tuberculosis are evolutionarily hyperconserved. Nat. Genet. 42, 498–503 (2010).

36. Correa-Macedo, W., Cambri, G. & Schurr, E. The Interplay of Human and Mycobacterium Tuberculosis Genomic Variability. Front. Genet. 10, (2019).

37. Salie, M. et al. Associations between human leukocyte antigen class I variants and the Mycobacterium tuberculosis subtypes causing disease. J. Infect. Dis. 209, 216–223 (2014).

38. Comas, I. et al. Out-of-Africa migration and Neolithic coexpansion of Mycobacterium tuberculosis with modern humans. Nat. Genet. 45, 1176–1182 (2013).

39. Coscolla, M. & Gagneux, S. Consequences of genomic diversity in Mycobacterium tuberculosis. Semin. Immunol. 26, 431–444 (2014).

40. Copin, R. et al. Within Host Evolution Selects for a Dominant Genotype of Mycobacterium tuberculosis while T Cells Increase Pathogen Genetic Diversity. PLoS Pathog. 12, e1006111 (2016).

41. Fox, G. J., Orlova, M. & Schurr, E. Tuberculosis in Newborns: The Lessons of the “Lübeck Disaster” (1929–1933). PLoS Pathog. 12, (2016).

42. Demerath, E. W. et al. Genome-wide association study of age at menarche in African-American women. Hum. Mol. Genet. 22, 3329–3346 (2013).

43. Elks, C. E. et al. Thirty new loci for age at menarche identified by a meta-analysis of genome-wide association studies. Nat. Genet. 42, 1077–1085 (2010).

44. Colas, E. et al. Molecular markers of endometrial carcinoma detected in uterine aspirates. Int. J. Cancer 129, 2435–2444 (2011).

45. Wu, Y. et al. Germline mutations in PPFIBP2 are associated with lethal prostate cancer. The Prostate 78, 1222–1228 (2018).

46. Messina, G. et al. The human Cranio Facial Development Protein 1 (Cfdp1) gene encodes a protein required for the maintenance of higher-order chromatin organization. Sci. Rep. 7, 45022 (2017).

47. Daya, M. et al. A Panel of Ancestry Informative Markers for the Complex Five-Way Admixed South African Coloured Population. PLOS ONE 8, e82224 (2013).

48. Brown, M. T. et al. ASAP1, a phospholipid-dependent arf GTPase-activating protein that associates with and is phosphorylated by Src. Mol. Cell. Biol. 18, 7038–7051 (1998).

49. Daya, M., van der Merwe, L., van Helden, P. D., Möller, M. & Hoal, E. G. The role of ancestry in TB susceptibility of an admixed South African population. Tuberc. Edinb. Scotl. 94, 413– 420 (2014).

50. Chen, C. et al. A Common Variant of ASAP1 Is Associated with Tuberculosis Susceptibility in the Han Chinese Population. Dis. Markers 2019, 7945429 (2019).

51. Wang, X., Ma, A., Han, X., Litifu, A. & Xue, F. ASAP1 gene polymorphisms are associated with susceptibility to tuberculosis in a Chinese Xinjiang Muslim population. Exp. Ther. Med. 15, 3392–3398 (2018).

52. Hoal, E. G., Dippenaar, A., Kinnear, C., van Helden, P. D. & Möller, M. The arms race between man and Mycobacterium tuberculosis: Time to regroup. Infect. Genet. Evol. J. Mol. Epidemiol. Evol. Genet. Infect. Dis. (2017) doi:10.1016/j.meegid.2017.08.021.

53. Campbell, M. C. & Tishkoff, S. A. African genetic diversity: implications for human demographic history, modern human origins, and complex disease mapping. Annu. Rev. Genomics Hum. Genet. 9, 403–433 (2008).

54. Peprah, E., Xu, H., Tekola-Ayele, F. & Royal, C. D. Genome-Wide Association Studies in Africans and African Americans: Expanding the Framework of the Genomics of Human Traits and Disease. Public Health Genomics 18, 40–51 (2015).

55. Simmons, J. D. et al. Immunological mechanisms of human resistance to persistent Mycobacterium tuberculosis infection. Nat. Rev. Immunol. 18, 575–589 (2018).

56. Munafò, M. R., Tilling, K., Taylor, A. E., Evans, D. M. & Davey Smith, G. Collider scope: when selection bias can substantially influence observed associations. Int. J. Epidemiol. 47, 226– 235 (2018).

57. Chang, C. C. et al. Second-generation PLINK: rising to the challenge of larger and richer datasets. GigaScience 4, 7 (2015).

58. Delaneau, O., Zagury, J.-F. & Marchini, J. Improved whole-chromosome phasing for disease and population genetic studies. Nat. Methods 10, 5–6 (2013).

59. Howie, B. N., Donnelly, P. & Marchini, J. A flexible and accurate genotype imputation method for the next generation of genome-wide association studies. PLoS Genet. 5, e1000529 (2009).

60. Sudmant, P. H. et al. An integrated map of structural variation in 2,504 human genomes. Nature 526, 75–81 (2015).

61. Schurz, H. et al. Evaluating the Accuracy of Imputation Methods in a Five-Way Admixed Population. Front. Genet. 10, (2019).

62. Deelen, P. et al. Genotype harmonizer: automatic strand alignment and format conversion for genotype data integration. BMC Res. Notes 7, 901 (2014).

63. Kent, W. J. et al. The Human Genome Browser at UCSC. Genome Res. 12, 996–1006 (2002).

64. Yang, J., Lee, S. H., Goddard, M. E. & Visscher, P. M. GCTA: A Tool for Genome-wide Complex Trait Analysis. Am. J. Hum. Genet. 88, 76–82 (2011).

65. Mägi, R. et al. Trans-ethnic meta-regression of genome-wide association studies accounting for ancestry increases power for discovery and improves fine-mapping resolution. Hum. Mol. Genet. 26, 3639–3650 (2017).

66. Mägi, R. & Morris, A. P. GWAMA: software for genome-wide association meta-analysis. BMC Bioinformatics 11, 288 (2010).

67. Panagiotou, O. A., Ioannidis, J. P. A., & for the Genome-Wide Significance Project. What should the genome-wide significance threshold be? Empirical replication of borderline genetic associations. Int. J. Epidemiol. 41, 273–286 (2012).

68. R Development Core Team. R: A language and environment for statistical computing. http://www.r-project.org. (2013).

69. Zheng, X. Imputation-Based HLA Typing with SNPs in GWAS Studies. in HLA Typing: Methods and Protocols (ed. Boegel, S.) 163–176 (Springer, 2018). doi:10.1007/978-1-4939-8546-3_11.

70. Zheng, X. et al. HIBAG—HLA genotype imputation with attribute bagging. Pharmacogenomics J. 14, 192–200 (2014).

71. Viechtbauer, W. & others. Conducting meta-analyses in R with the metafor package. J. Stat. Softw. 36, 1–48 (2010).

72. Satorra, A. & Bentler, P. M. A scaled difference chi-square test statistic for moment structure analysis. Psychometrika 66, 507–514 (2001).

73. Mahajan, A. et al. Genome-wide trans-ancestry meta-analysis provides insight into the genetic architecture of type 2 diabetes susceptibility. Nat. Genet. 46, 234–244 (2014).

