## Supplementary Tables and Figures for "Multi-ancestry meta-analysis of host genetic susceptibility to tuberculosis identifies shared genetic architecture"

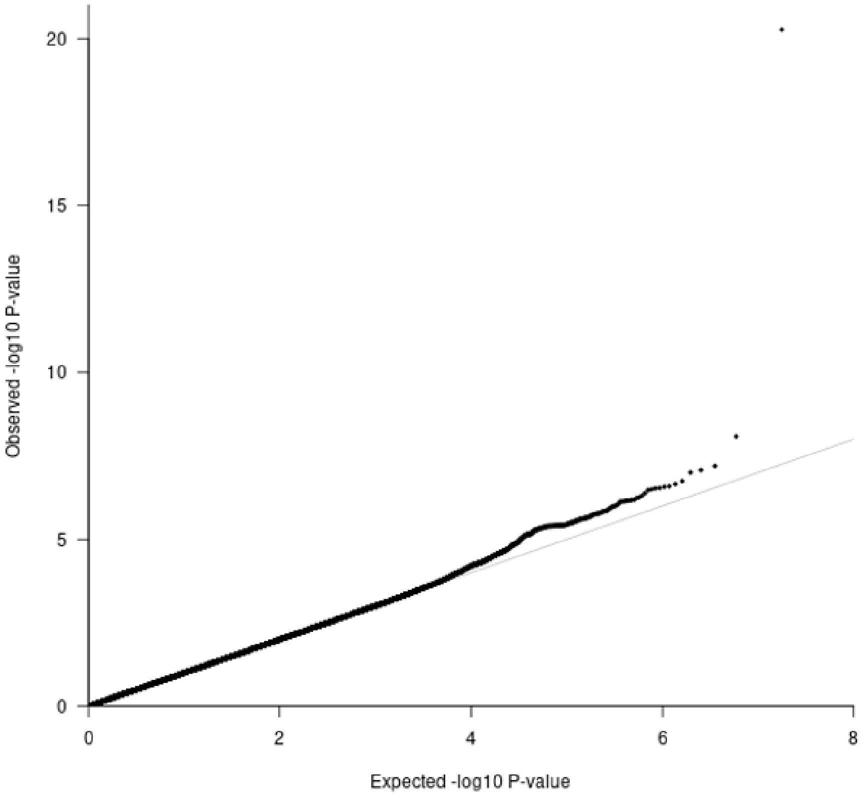


| Study | Lambda |
|---|---|
| China1 | 0.93 |
| China2 | 1.00 |
| China3 | 1.01 |
| Japan | 0.99 |
| Thailand | 1.00 |
| Russia | 1.00 |
| Estonia | 1.00 |
| Germany | 1.00 |
| Gambia | 1.00 |
| Ghana | 1.00 |
| SAC_A | 1.03 |
| SAC_M | 1.00 |

**Figure S1**: QQ-plot (left) from the MR-MEGA analysis of all 12 datasets with genomic control correction, including two PCs as covariates. The lambda value of 1.00 suggests no significant inflation or deflation of p-values. Lambda values for the input data (right).

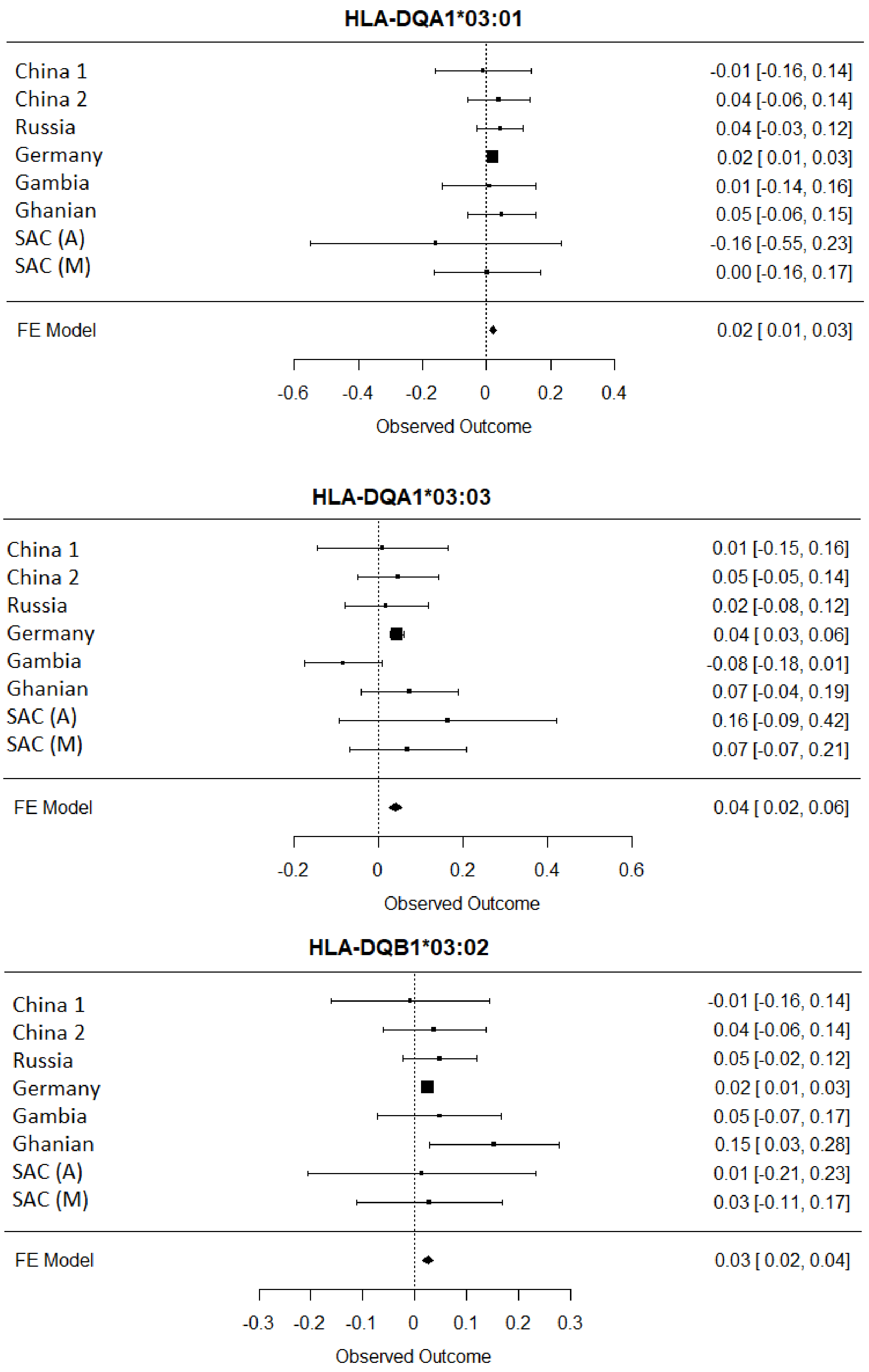


**Figure S2:** Trans ethnic meta-analysis results for the three replicated HLA epitope association, showing consistent direction of genetic effects between the input studies.

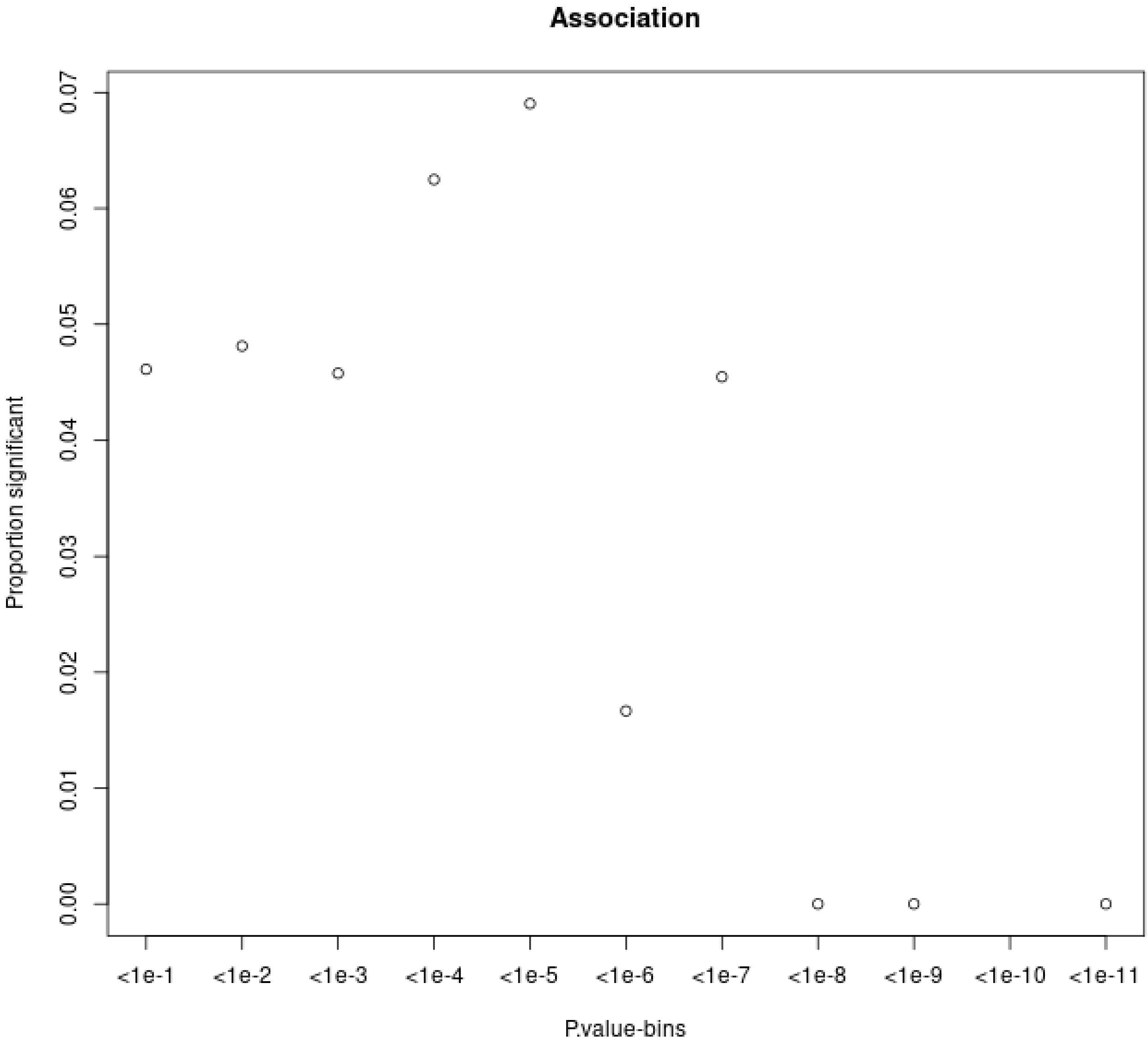


**Figure S3**: Proportion of variants that had a significant change in association p-value following the inclusion of the force of infection p-value for different p-value bins.

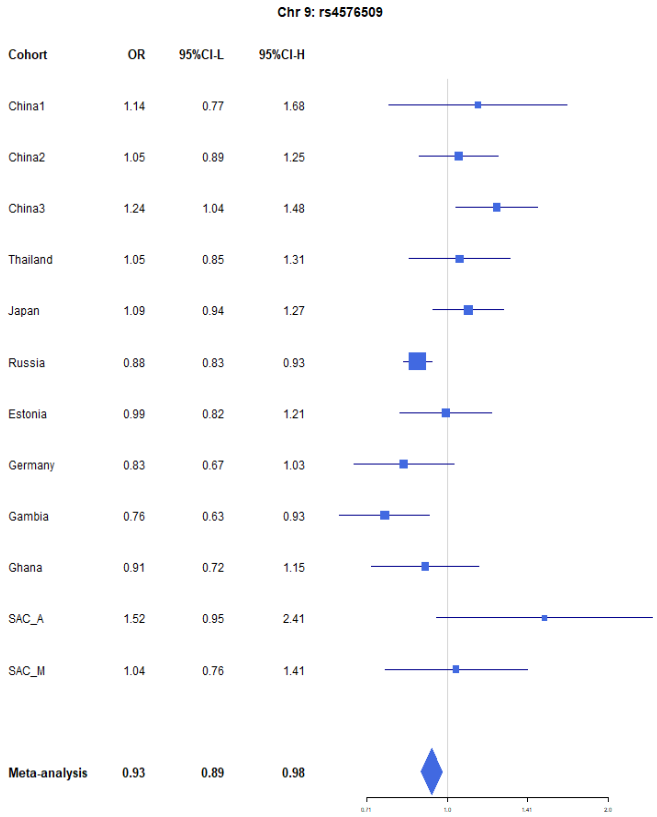


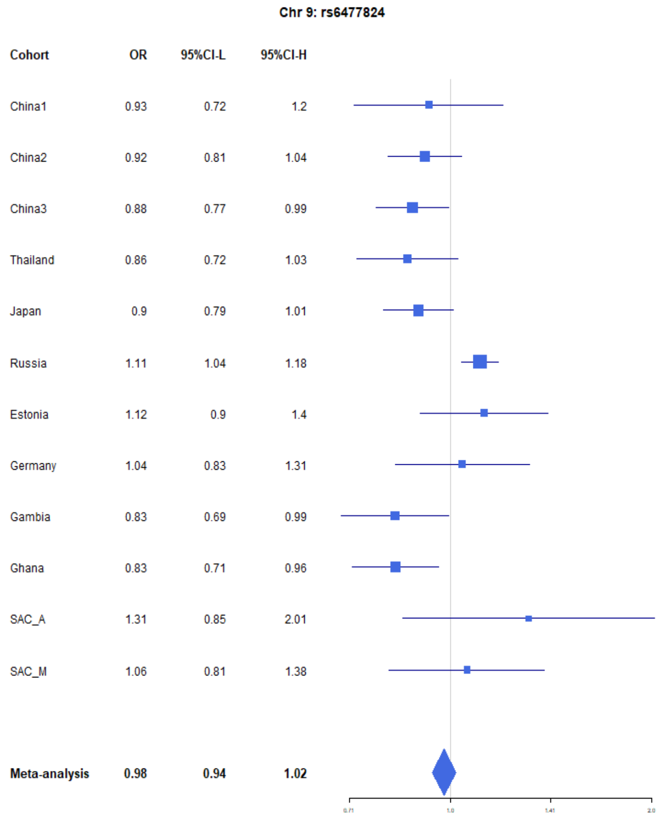


**Figure S4:** Forest plots for the suggestive chromosome 9 peaks, rs4576509 (left) and rs6477824 (right) for the trans-ethnic Mr-MEGA analysis including all 12 cohorts.

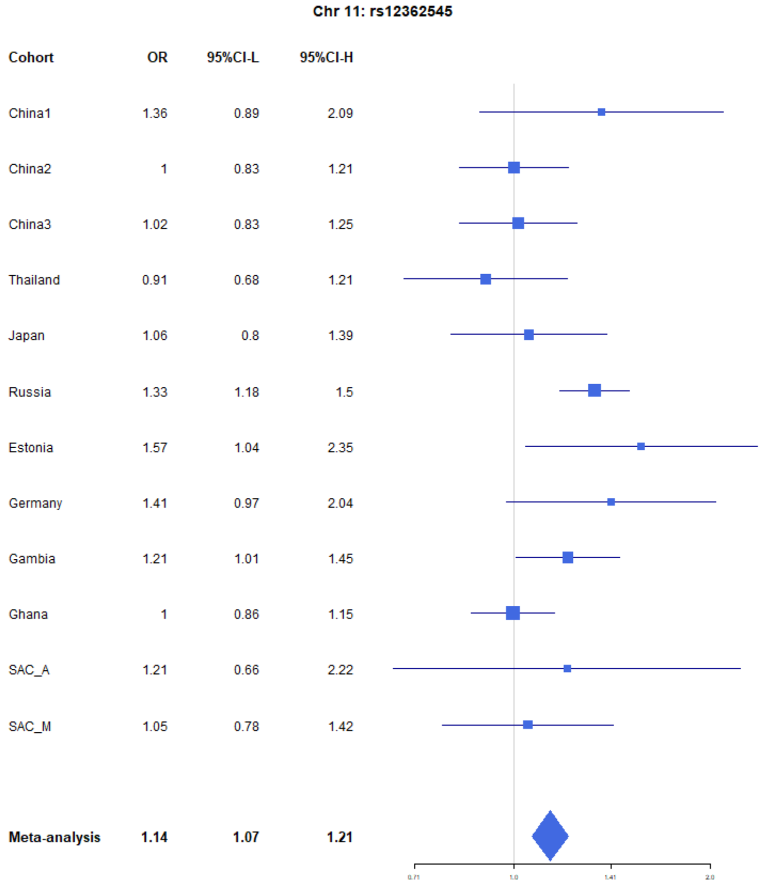


**Figure S5:** Forest plots for the suggestive chromosome 11 peak, rs12362545, for the trans-ethnic Mr-MEGA analysis including all 12 cohorts.

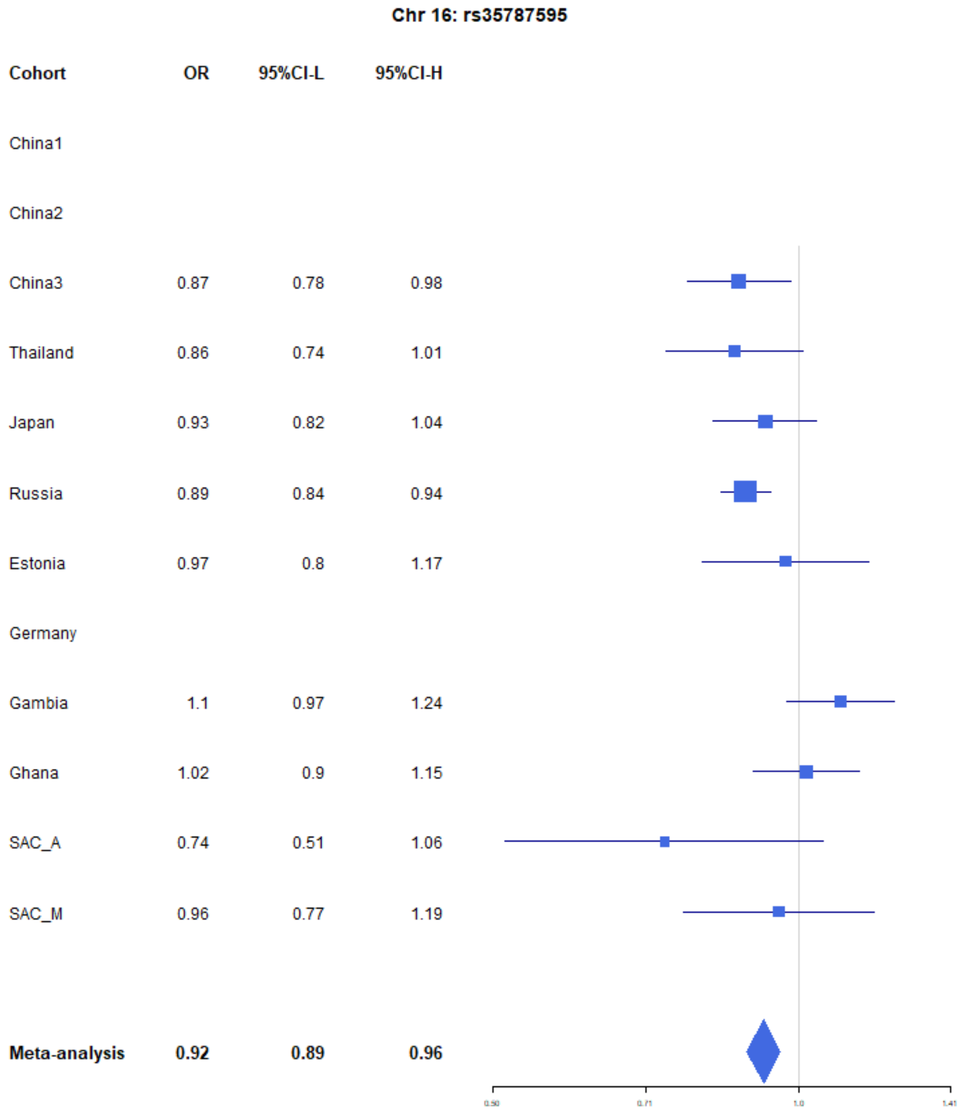


**Figure S6:** Forest plots for the suggestive chromosome 16 peak, rs35787595, for the trans-ethnic Mr-MEGA analysis including all 12 cohorts.

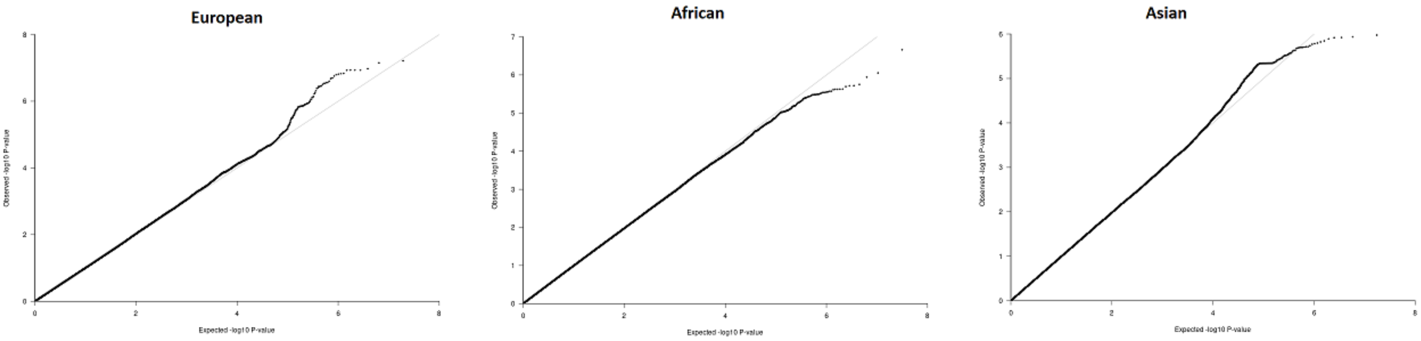


**Figure S7:** QQ-plots for the region-specific FE meta-analysis using GCC and implemented in GWAMA

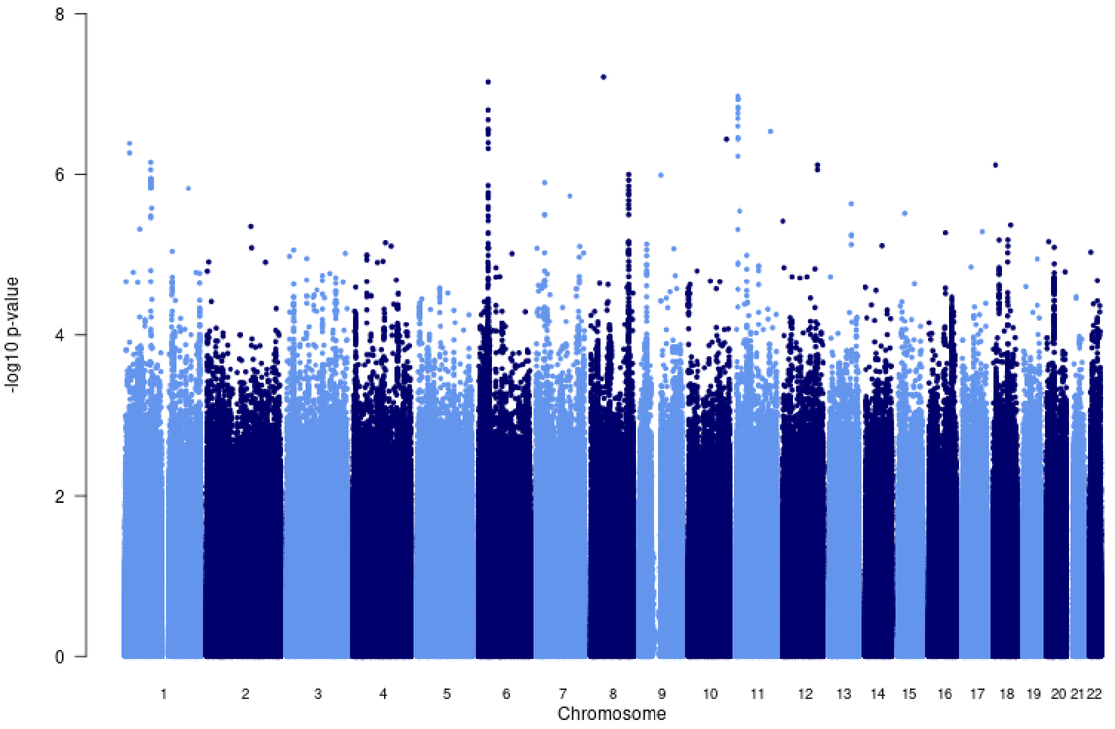


**Figure S8:** Manhattan plot of all p-values (>=2studies) for the European subgroup analysis. FE model with GCC implemented in GWAMA.

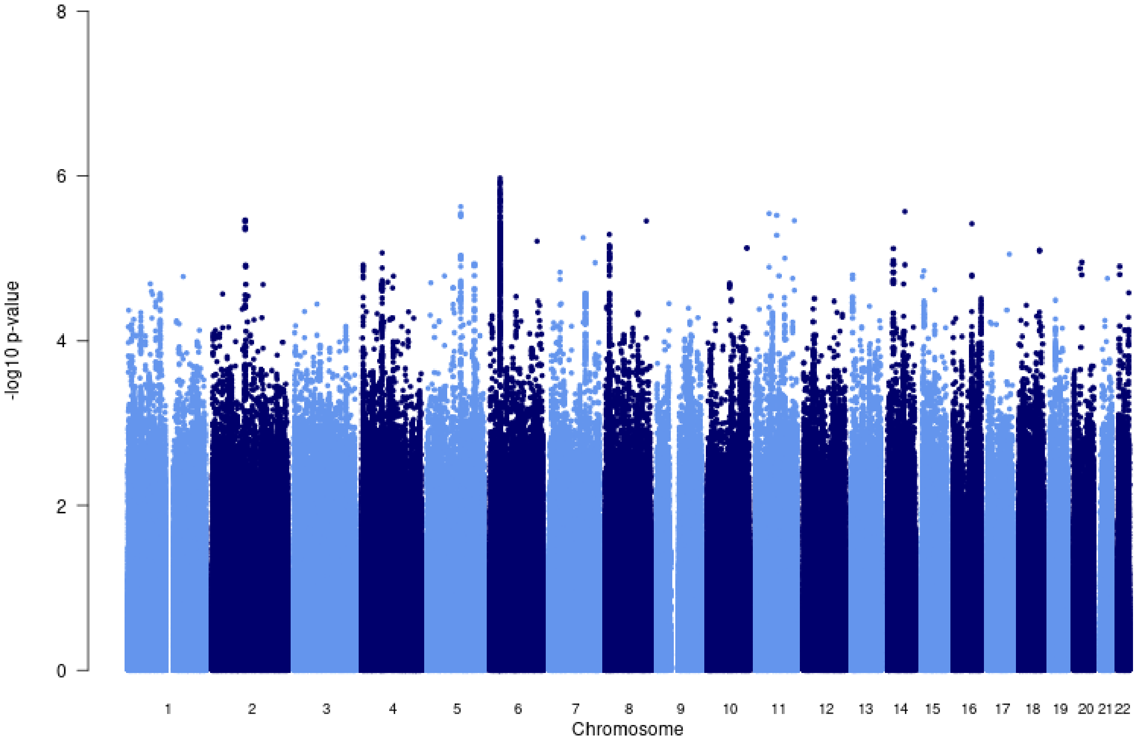


**Figure S9:** Manhattan plot of all p-values (>=2studies) for the Asian subgroup analysis. FE model with GCC implemented in GWAMA.

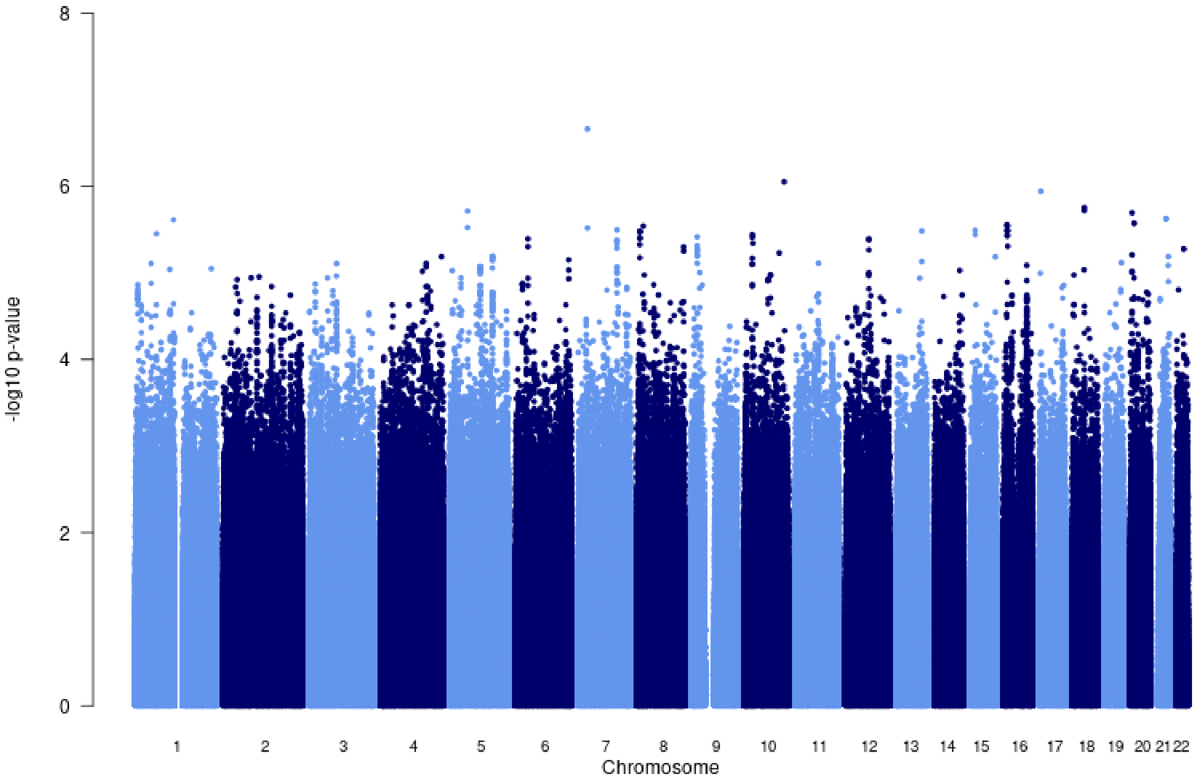


**Figure S10:** Manhattan plot of all p-values (>=2studies) for the African subgroup analysis. FE model with GCC implemented in GWAMA.

**Table S1**: Summary of ITHGC TB-GWAS datasets.

| **Dataset** | **Population** | **Cases/ Controls** | **Control definition** | **TB diagnosis** | **TB prevalence per 100 000 pa.** | **Estimated proportion of controls ever exposed to *Mtb* (±SD)**** | **#SNPs** | **Genotyping platform** | **Reference** |
|---|---|---|---|---|---|---|---|---|---|
| **China 1*** | Asian | 483/ 587 | A peer-nominated approach was used to recruit control subjects to the study.*** | Acid-fast staining and culturing of Mtb from sputum samples. | 89 | 0.302 (0.101) | 7 710 153 | Affymetrix Genome-Wide Human SNP Array 6.0 | (unpublished) |
| **China 2*** | Asian | 1290/ 1145 | A peer-nominated approach was used to recruit control subjects to the study.*** | Acid-fast staining and culturing of Mtb from sputum samples. | 89 | 0.302 (0.101) | 9 769 029 | Illumina Human OmniZhonghua-8 chips. | (unpublished) |
| **China 3** | Asian | 972/ 1537 | Controls were defined as individuals with no history of TB, normal radiographic findings, and negative purified protein de- | Acid-fast staining and culturing of Mtb from sputum samples. | 89 | 0.302 (0.101) | 9 726 450 | Illumina Human OmniZhonghua-8 chips. | (Qi et al., 2017) |

| Dataset | Population | Cases/ Controls | Control definition | TB diagnosis | TB prevalence per 100 000 pa. | Estimated proportion of controls ever exposed to *Mtb* (±SD)** | #SNPs | Genotyping platform | Reference |
|---|---|---|---|---|---|---|---|---|---|
| | | | rivative (PPD) skin tests (<5 mm) | | | | | | |
| **Thailand** | Asian | 433/ 295 | Blood donors screened for a history of TB and a family history of TB | Acid-fast staining and culturing of Mtb from sputum samples. | 236 | 0.404 (0.112) | 6 723 358 | Illumina Human610-Quad | (Mahasirimongkol et al., 2012) |
| **Japan** | Asian | 751/ 3199 | Blood donors screened for a history of TB and a family history of TB | Acid-fast staining and culturing of Mtb from sputum samples. | 23 | 0.142 (0.125) | 9 051 051 | Illumina HumanHap550 | (Mahasirimongkol et al., 2012) |
| **Russia*** | European | 5914/ 6022 | Healthy adult blood bank donors with no history of TB. *Mtb* infection status of these controls was unknown | Acid-fast staining and culturing of Mtb from sputum samples. | 109 | 0.191 (0.093) | 10 878 777 | Affymetrix Genome-Wide Human SNP Array 6.0 | (Curtis et al., 2015) |
| **Estonia** | European | 62/ 2660 | Healthy Biobank controls | Biobank TB cases | 13 | 0.116 (0.093) | 10 611 556 | Illumina 370K | |

| Dataset | Population | Cases/ Controls | Control definition | TB diagnosis | TB prevalence per 100 000 pa. | Estimated proportion of controls ever exposed to *Mtb* (±SD)** | #SNPs | Genotyping platform | Reference |
|---|---|---|---|---|---|---|---|---|---|
| | | | | | | | | | (unpublished) |
| **Germany*** | European | 586/ 333 | A peer-nominated approach was used to recruit control subjects to the study.*** | NA | 7.8 | 0.067 (0.081) | 10 602 193 | Illumina Omni2.5 +exome | (unpublished) |
| **Gambia*** | African | 1316/ 1382 | A peer-nominated approach was used to recruit control subjects to the study.*** | NA | 126 | 0.280 (0.089) | 18 634 017 | Affymetrix GeneChip 500K | (Consortium, 2007) |
| **Ghana*** | African | 1359/ 1952 | A peer-nominated approach was used to recruit control subjects to the study.*** | Acid-fast staining and culturing of Mtb from sputum samples. | 282 | 0.539 (0.198) | 19 029 214 | Affymetrix Genome-Wide Human SNP Array 6.0 | (Thye et al., 2010) |
| **SAC(A)* ^** | African | 577/19 | Community contacts assumed to be exposed | Acid-fast staining and culturing of Mtb from | 717 | 0.436 (0.127) | 9 227 330 | Affymetrix 500k | (Daya et al., 2014b) |

| Dataset | Population | Cases/ Controls | Control definition | TB diagnosis | TB prevalence per 100 000 pa. | Estimated proportion of controls ever exposed to *Mtb* (±SD)** | #SNPs | Genotyping platform | Reference |
|---|---|---|---|---|---|---|---|---|---|
| | | | and latently infected with Mtb | sputum samples. | | | | | |
| **SAC(M) * ^** | African | 410/ 405 | Community contacts assumed to be exposed and latently infected with Mtb | Acid-fast staining and culturing of Mtb from sputum samples. | 717 | 0.436 (0.127) | 11 371 838 | Illumina MEGA array | (Schurz et al., 2019a) |

* Raw genotyping data available

** Estimated proportion of control individuals ever infected with Mtb by age 35-44 in 2010, based on data from Houben & Dodd

^ SAC(A/M): South African admixed population (SAC) Affymetrix (A) and MEGA (M) array data

*** With this approach, patient participants were asked to take an information sheet home and to invite friends/neighbours to participate in the study. It was made clear that the invitee should have no history of TB, have no symptoms of TB (e.g. cough), be of similar age, sex and ethnicity and, if they agree to participate, they should join the patient on his/her next clinical follow-up. At the clinic, it was emphasized again that the participant is under no obligation to take part. They were informed as to the nature and scope of the project and their role and rights as a participant prior to providing written consent. For all participants, age, sex, ethnicity, education, occupation, and TB vaccination data were collected in a short questionnaire.

**Table S2:** Polygenic heritability estimates at different TB prevalence rates.

| Cohort | Prevalence | VG (SE)* | V(G)/Vp_L (SE)* | | |
|---|---|---|---|---|---|
| | | | 0.1x' | 1x ' | 10x ' |
| **China1** | 0.00089 | 0.125 (0.045) | 0.195 (0.069) | 0.349 (0.124) | 0.542 (0.192) |
| **China2** | 0.00089 | 0.149 (0.037) | 0.116 (0.028) | 0.208 (0.050) | 0.324 (0.078) |
| **China3** | 0.00088 | 0.134 (0.037) | 0.119 (0.032) | 0.216 (0.059) | 0.336 (0.091) |
| **Japan** | 0.00023 | 0.027 (0.014) | 0.012 (0.006) | 0.083 (0.043) | 0.119 (0.061) |
| **Thailand** | 0.00236 | 0.029 (0.024) | 0.036 (0.029) | 0.051 (0.042) | 0.087 (0.070) |
| **Russia** | 0.00109 | 0.411 (0.014) | 0.219 (0.005) | 0.357 (0.008) | 0.563 (0.013) |
| **Germany** | 0.00008 | 0.231 (0.054) | 0.007 (0.001) | 0.189 (0.041) | 0.366 (0.080) |
| **Gambia** | 0.00126 | 0.304 (0.033) | 0.235 (0.023) | 0.366 (0.036) | 0.583 (0.058) |
| **Ghana** | 0.00282 | 0.091 (0.034) | 0.116 (0.043) | 0.168 (0.063) | 0.286 (0.107) |
| **SAC** | 0.00717 | 0.151 (0.096) | 0.196 (0.123) | 0.300 (0.188) | 0.556 (0.348) |
| **Average** | NA | 0.240 (0.009) | 0.153 (0.007) | 0.263 (0.013) | 0.419 (0.022) |

* VG (SE): Genetic variance estimate and standard error
*V(G)/Vp_L (SE): Ratio of genetic variance to phenotypic variance, estimate and standard error
'Prevalence multiplier

**Table S3:** Suggestive associations (p-value <= $1e^{-5}$) for the multi-ancestry analysis including data from all 12 datasets implementing MR-Mega analysis with GCC.

| Marker Name | Chromosome | Position | Gene | Location | CADD score | EA ^ | NEA ^ | EAF ^ | Sample size | Datasets | P-value |
|---|---|---|---|---|---|---|---|---|---|---|---|
| **rs28578990** | 6 | 32505643 | *HLA-DRB5* | intergenic | 1.766 | G | A | 0.037 | 24378 | 7 | $2.90e^{-07}$ |
| **rs114803904** | 6 | 32694001 | *HLA-DQB3* | intergenic | 0.78 | C | T | 0.393 | 24377 | 7 | $4.88e^{-06}$ |
| **rs111718686** | 6 | 32697405 | *HLA-DQB3* | intergenic | 12.57 | C | T | 0.169 | 18205 | 5 | $2.72e^{-06}$ |
| **rs2621322** | 6 | 32788712 | *TAP2* | intronic | 0.27 | G | T | 0.231 | 26359 | 10 | $4.52e^{-06}$ |
| **rs146049519** | 6 | 32555142 | *HLA-DRB1* | intronic | 0.1 | C | G | 0.035 | 24377 | 7 | $1.03e^{-06}$ |
| **rs73409538** | 6 | 32611624 | *HLA-DQA1* | downstream | 12.51 | T | G | 0.035 | 24377 | 7 | $1.52e^{-06}$ |
| **rs28383323** | 6 | 32594039 | *HLA-DQA1* | intergenic | 4.47 | A | G | 0.260 | 26330 | 10 | $6.36e^{-08}$ |
| **rs28680981** | 6 | 32606579 | *HLA-DQA1* | intronic | 12.23 | A | G | 0.232 | 25584 | 9 | $1.01e^{-07}$ |
| **rs28606662** | 6 | 32614864 | *HLA-DQA1* | intergenic | 5.72 | G | C | 0.197 | 25550 | 9 | $2.84e^{-06}$ |
| **rs112984211** | 6 | 32617889 | *HLA-DQA1* | intergenic | 5.57 | T | G | 0.184 | 24378 | 7 | $2.14e^{-06}$ |

| Marker Name | Chromosome | Position | Gene | Location | CADD score | EA ^ | NEA ^ | EAF ^ | Sample size | Datasets | P-value |
|---|---|---|---|---|---|---|---|---|---|---|---|
| **rs2858331** | 6 | 32681277 | *XXbac-BPG254F23.7* | intergenic | 2.49 | G | A | 0.482 | 26352 | 10 | 7.02 $e^{-07}$ |
| **rs6913309** | 6 | 32339840 | *C6orf10* | upstream | 3.55 | A | T | 0.274 | 26330 | 10 | 3.18 $e^{-07}$ |
| **rs80234155** | 6 | 32512483 | *RNU1-61P* | intergenic | 2.83 | C | G | 0.169 | 25110 | 8 | 2.21 $e^{-07}$ |
| **rs115752743** | 6 | 32559163 | *HLA-DRB1* | intergenic | 6.93 | T | C | 0.228 | 25102 | 8 | 2.67 $e^{-07}$ |
| **rs6477824** | 9 | 114287453 | *ZNF483* | UTR5 | 9.43 | C | T | 0.745 | 31279 | 10 | 2.99 $e^{-07}$ |
| **rs4576509** | 9 | 23294037 | *SUMO2P2* | intergenic | 16.2 | G | C | 0.768 | 31224 | 10 | 7.40 $e^{-07}$ |
| **rs12362545** | 11 | 7573173 | *PPFIBP2* | intronic | 5.167 | T | G | 0.078 | 32633 | 12 | 1.24 $e^{-06}$ |
| **rs35787595** | 16 | 75466847 | *CFDP1* | intronic | 4.5 | C | G | 0.511 | 28299 | 9 | 5.41 $e^{-06}$ |

**Table S4:** Results for the concordance in direction of effect analysis for alp-value thresholds and reference populations (for SNNP selection)

| Reference | Threshold | Comparison | p-value | probability of success |
|---|---|---|---|---|
| Europe | p-value <= 0.001 | Europe vs. Africa | 0.477 | 0.503 |
| | | Europe vs. Asia | 0.831 | 0.474 |
| | 0.001< p-value <= 0.01 | Europe vs. Africa | 0.00614 | 0.531 |
| | | Europe vs. Asia | 0.2843 | 0.507 |
| | 0.01< p-value <= 0.5 | Europe vs. Africa | 0.220 | 0.505 |
| | | Europe vs. Asia | 0.650 | 0.497 |
| | 0.5< p-value <= 1.0 | Europe vs. Africa | 0.286 | 0.504 |
| | | Europe vs. Asia | 0.558 | 0.499 |
| Asia | p-value <= 0.001 | Asia vs. Europe | 0.293 | 0.518 |
| | | Asia vs. Africa | 0.707 | 0.485 |
| | 0.001< p-value <= 0.01 | Asia vs. Europe | 0.698 | 0.493 |
| | | Asia vs. Africa | 0.68 | 0.494 |
| | 0.01< p-value <= 0.5 | Asia vs. Europe | 0.599 | 0.498 |
| | | Asia vs. Africa | 0.541 | 0.499 |
| | 0.5< p-value <= 1.0 | Asia vs. Europe | 0.271 | 0.504 |
| | | Asia vs. Africa | 0.482 | 0.500 |
| Africa | p-value <= 0.001 | Africa vs. Europe | 0.432 | 0.504 |
| | | Africa vs. Asia | 0.885 | 0.475 |
| | 0.001< p-value <= 0.01 | Africa vs. Europe | 0.628 | 0.496 |
| | | Africa vs. Asia | 0.418 | 0.502 |
| | 0.01< p-value <= 0.5 | Africa vs. Europe | 0.091 | 0.509 |
| | | Africa vs. Asia | 0.75 | 0.495 |
| | 0.5< p-value <= 1.0 | Africa vs. Europe | 0.014 | 0.516 |
| | | Africa vs. Asia | 0.627 | 0.497 |

**Table S5:** Suggestive associations for the European and Asian ancestry-specific FE analysis (with GCC)

| **Rs number (chr)** | **EA^** | **NEA^** | **EAF^** | **Gene** | **OR** | **OR 95L*** | **OR 95U*** | **p-value** | **Datasets** | **Sample size** | **Effects**** | **Pop** |
|---|---|---|---|---|---|---|---|---|---|---|---|---|
| **rs28383206 (6)** | G | A | 0.20 | HLA | 0.82 | 0.77 | 0.88 | 7.06e-08 | 3 | 14791 | --+ | European |
| **rs3935174 (8)** | T | A | 0.30 | ASAP1 | 0.87 | 0.82 | 0.92 | 1.00e-06 | 3 | 14791 | -+- | European |
| **rs12362545 (11)** | T | G | 0.05 | PFIBP2 | 1.35 | 1.21 | 1.51 | 1.06e-07 | 3 | 14791 | +++ | European |
| **rs146049519 (6)** | C | G | 0.032 | HLA-DRB1 | 2.1 | 1.59 | 2.95 | 1.06e-06 | 2 | 3578 | ??+?+ | Asian |
| **rs62495207 (8)** | C | G | 0.46 | | 0.83 | 0.77 | 0.904 | 5.10e-06 | 3 | 7174 | --??- | Asian |

* Lower (L) and upper (U) 95% confidence interval of the odds ratio (OR)

** Effect (OR) direction of individual input studies, either negative (-) positive (+) or not present (?)

^EA and EAF: Effect allele and effect allele frequency

^NEA: Non-effect allele
